# An individual participant data meta-analysis of prognostic blood biomarkers in IPF

**DOI:** 10.1101/2021.03.30.21254621

**Authors:** Fasihul A Khan, Iain Stewart, Gauri Saini, Karen A. Robinson, R Gisli Jenkins

## Abstract

**Background:** Idiopathic pulmonary fibrosis (IPF) is a progressive fibrotic lung disease with variable disease trajectory. Blood biomarkers reflecting disease severity that can accurately predict outcomes are urgently needed. Through systematic review and meta-analysis, we evaluate the prognostic potential of matrix-metalloproteinase-7 (MMP-7) and other frequently studied blood biomarkers in patients with IPF.

**Methods:** Electronic databases were searched on 12^th^ November 2020 to identify prospective studies reporting outcomes in patients with untreated IPF, stratified according to at least one pre-specified biomarker, measured at either baseline or change over three months. Individual participant data (IPD) was sought for studies investigating MMP-7 as a prognostic factor. The primary outcome was overall mortality, with secondary outcomes including disease progression, defined as >10% relative FVC decline or death.

**Results:** 29 studies reporting outcomes from 3950 IPF participants were included, investigating a total of 16 biomarkers. IPD from MMP-7 studies was available for eleven cohorts. Baseline MMP-7 levels were associated with increased mortality (adjusted HR1.23 per SD increase, 95%CI 1.03;1.48, I^2^=64.3%) and disease progression (adjusted OR1.27 per SD increase, 95%CI 1.11;1.46, I^2^=5.9%), but change in MMP-7 over three-months was not associated with any of the measured outcomes. There was insufficient data for quantitative analysis in non-MMP7 studies, and whilst many biomarkers showed an association with clinical outcomes, replication of effects across studies was weak.

**Conclusion:** Baseline MMP-7 levels were associated with an increased risk of overall mortality and disease progression in patients with untreated IPF. The evidence for other biomarkers is currently insufficient with further studies needed.

## INTRODUCTION

Idiopathic paulmonary fibrosis (IPF) is a chronic progressive fibrotic lung disease of unknown aetiology that affects approximately 3 million people worldwide, with a rising incidence and a median survival from diagnosis of approximately three years^1–5^. Disease trajectory is variable, ranging from slow progression to rapid loss of lung function and death^6^. The most recognised biomarker of disease progression in IPF is the change in forced vital capacity (FVC) at 12 months^7 8^. However, lung function measurements have limitations, including test variability related to patient effort and confounding effects of comorbidities such as emphysema.^9^

Blood proteomic biomarkers measured at diagnosis may have the potential to accurately judge disease severity and predict prognosis at an earlier timepoint, thus enabling a personalised approach to further management and modelling longitudinal disease behaviour^10^. Furthermore, short-term biomarkers of disease activity have the potential to transform early-phase clinical trials by acting as surrogate endpoints.

In IPF, biomarkers can be categorised according to possible pathogenic pathways, broadly including those associated with alveolar epithelial cell dysfunction, extracellular matrix (ECM) remodelling and fibroproliferation, as well as immune dysregulation. Whilst several blood biomarkers have been explored with respect to predicting prognosis in IPF, no published study has systematically synthesised this evidence. Through systematic review and meta-analysis of individual participant data (IPD), we explore the association between an epithelial biomarker, matrix-metalloproteinase-7 (MMP-7), and clinical endpoints in patients with untreated IPF. Simultaneously we evaluate the prognostic potential of other frequently studied blood biomarkers.

## METHODS

The systematic review was conducted in accordance with a pre-specified protocol (PROSPERO registration number: CRD42019120402) and has been reported using PRISMA-IPD (Preferred Reporting Items for Systematic Reviews and Meta-Analyses of Individual Participant Data) guidelines.^11^

### Search strategy and study selection

Electronic database searches were carried out in MEDLINE (1946 to latest), Embase (1974 to latest), Google Scholar, the Cochrane Register of Controlled Trials and ClinicalTrials.gov, with the last search carried out on 12^th^ November 2020. Keywords and controlled vocabulary terms for “idiopathic pulmonary fibrosis” and “biomarkers”, alongside search filters for prognostic studies were applied (Figure S1).^12^ Hand searches of reference lists in retrieved articles were conducted to identify further studies. Unpublished and ongoing studies were identified by searching pre-print servers including medRxiv, bioRxiv and Wellcome Open Research. Following searches, two reviewers independently in a standardised manner, screened through titles and abstracts before full text review. Disagreements were resolved by consensus, with unresolved conflicts decided by a third reviewer.

The review included all original prospective observational studies that reported outcomes in patients aged over 18 with anti-fibrotic naive IPF, diagnosed according to contemporaneous consensus guidelines,^13–15^ stratified according to at least one pre-identified blood biomarker. Conference abstracts reporting sufficient detail were eligible for inclusion. Retrospective studies, case reports, animal studies and studies investigating non-IPF interstitial lung disease (ILD) were excluded. Language or year of publication restrictions were not applied. No minimal study sample size was specified for inclusion.

Other than MMP-7, studies reporting the following biomarkers measured at either baseline and/or trends over 3 months were eligible: biomarkers of epithelial dysfunction [krebs von den Lungen-6 (KL-6), surfactant protein-A (SP-A), surfactant protein-D (SP-D), matrix metalloproteinase-1 (MMP-1), cancer antigen 125 (CA-125), carbohydrate antigen 19-9 (CA19-9), vascular endothelial growth factor (VEGF), insulin like growth factor binding protein 2 (IGFBP2)], biomarkers of ECM modelling [collagen synthesis peptides, neoepitopes, lysyl oxidase like 2 (LOXL2), periostin, osteopontin] and biomarkers of immune dysregulation [C-C motif chemokine ligand 18 (CCL-18), chemokine ligand 13 (CXCL13), interleukin-8 (IL-8), heat shock protein 70 (HSP70), chitinase-3-like protein 1 (YKL40), intracellular adhesion molecule 1 (ICAM-1)].

The primary outcome was overall mortality. Secondary outcomes measures included change in percent predicted FVC from baseline at 12 months and disease progression at 12 months defined as >10% FVC decline or death.

### Data extraction and risk of bias assessment

Data were extracted by one reviewer using an in-house proforma and verified by a second reviewer. Data included study design, participant and biomarkers characteristics, and outcome data including sample sizes, mean values and standard deviations of biomarkers in individuals with and without the event. Time to event data were collected using adjusted hazard ratios (HR) where reported.

IPD were sought from corresponding authors of studies investigating MMP-7 as a prognostic factor, using secure and encrypted electronic mail communication. A minimum of three reminders, each four weeks apart were sent. Data from sponsored clinical studies were requested through various online portals.^16–18^ Requested data included participant demographics (age, gender, smoking status and baseline lung function), baseline and three-month MMP-7 levels and outcomes including 12-month lung function and overall mortality (Figure S2). Where IPD were not made available, aggregate data were extracted from study publications.

Risk of bias assessment was carried out independently in duplicate using the Quality in Prognostic Studies (QUIPS) tool. The QUIPS tool^19^ assesses the risk of bias across six domains: study participation, study attrition, prognostic factor measurement, outcome measurement, study confounding and statistical analysis and reporting. All studies were included in the review irrespective of their risk of bias rating. The GRADE (Grading of Recommendations, Assessment, Development and Evaluations) framework was applied to rate the overall quality of evidence for each outcome as high, moderate, low or very low.^20^

### Statistical analysis

All identified studies were included in the data synthesis, with summary tables for study characteristics. Multiple cohorts within the same study were treated as individual cohorts. Due to methodological heterogeneity and marked difference in outcome measures, studies evaluating the role of biomarkers other than MMP-7 have been described narratively and in tables. Hazard ratios (HR) for MMP-7 levels in predicting mortality, and odds ratios (OR) for predicting disease progression, were estimated using a two-stage IPD meta-analysis with random effects and presented as forest plots. Estimates were adjusted for *a priori* confounders including age, sex, smoking history and baseline FVC. Studies with a follow up duration longer than three years were censored for survival analyses. Disease progression was dichotomised and defined as 10% relative decline in FVC or death within 12 months of baseline. To standardise biomarker values across studies, z scores specific to each study were calculated and analysed as exposure variables. The difference in MMP-7 over three-months was calculated using relative percent change from baseline. Participants with missing data were excluded using listwise deletion. The I^2^ statistic was used to evaluate statistical heterogeneity between studies. Meta-regression was conducted where sufficient studies were included to explore variability in heterogeneity owing to study design (cohort vs. randomised trial), single-centre studies, non-peer reviewed manuscripts, assay methods (ELISA vs. non-ELISA), and the type of blood samples used (serum vs. plasma). Publication bias was assessed using funnel plot analysis and Egger’s test.^21^ All statistical analyses were performed using Stata 16 (Statacorp, Texas US).

## RESULTS

Searches of the electronic databases on 12^th^ November 2020 yielded 4930 articles, with a further 69 studies identified through preprint servers. Following the removal of duplicates, screening and full text review, 29 studies published worldwide between 2007 and 2020, reporting outcomes from 3950 IPF participants were included (Figure 1). A total of 16 blood biomarkers were evaluated across the included studies. Individual study characteristics are presented in Table 1. A total of 12 studies reported outcomes in relation to MMP-7, of which IPD was available for nine studies (75%) reporting data from eleven individual cohorts and 1664 participants. No issues with the integrity of IPD were identified.

**Table 1.**
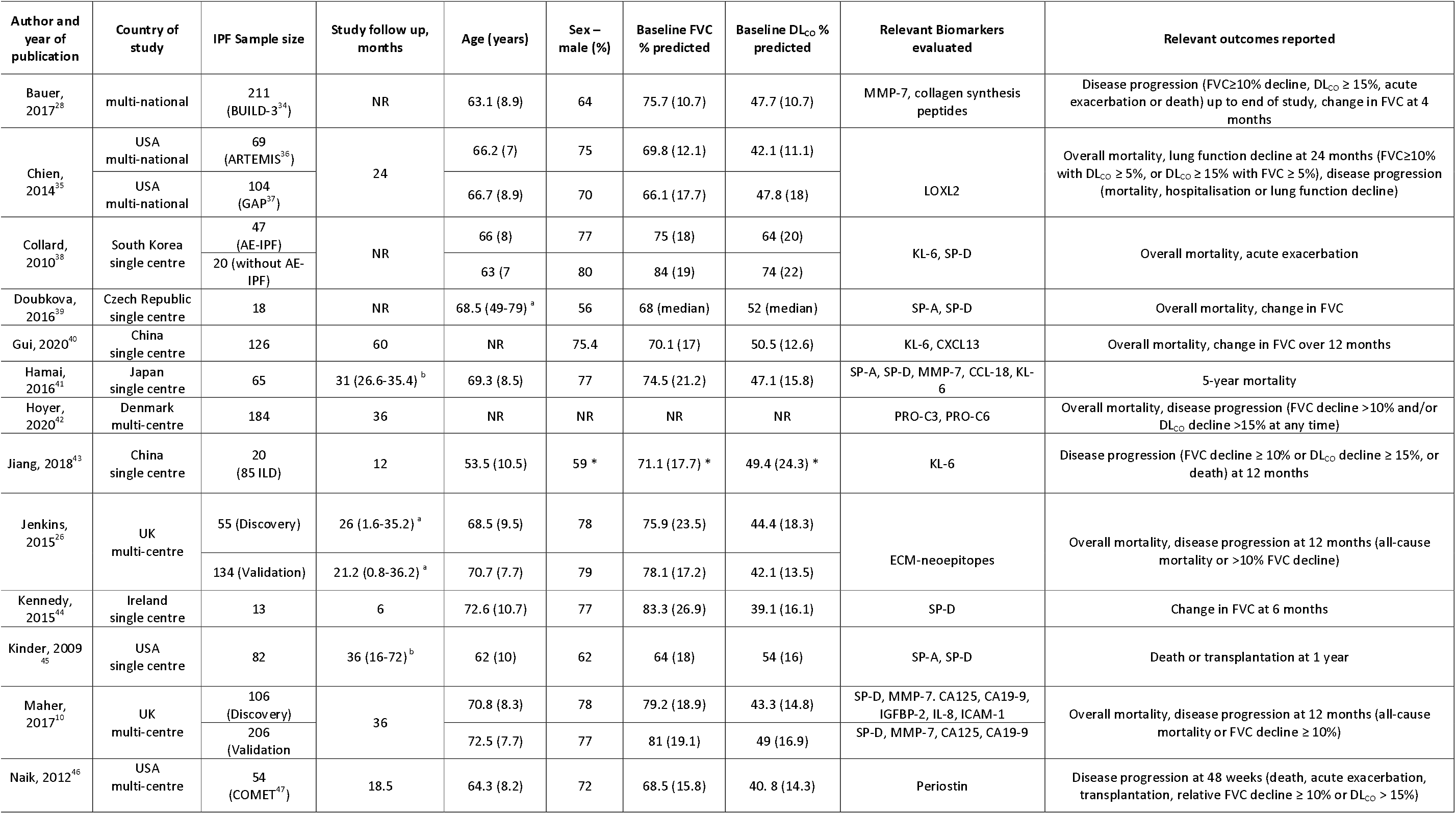
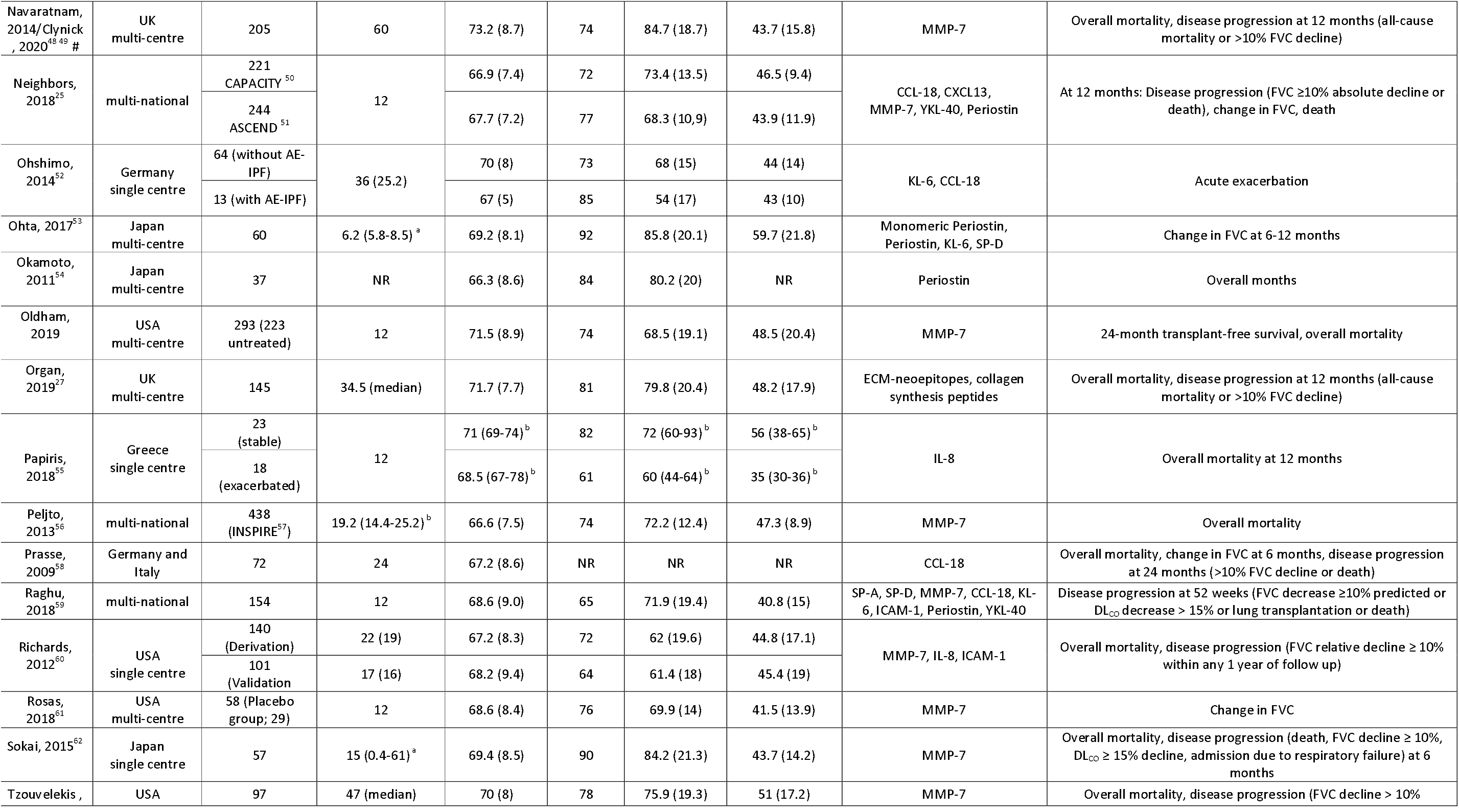
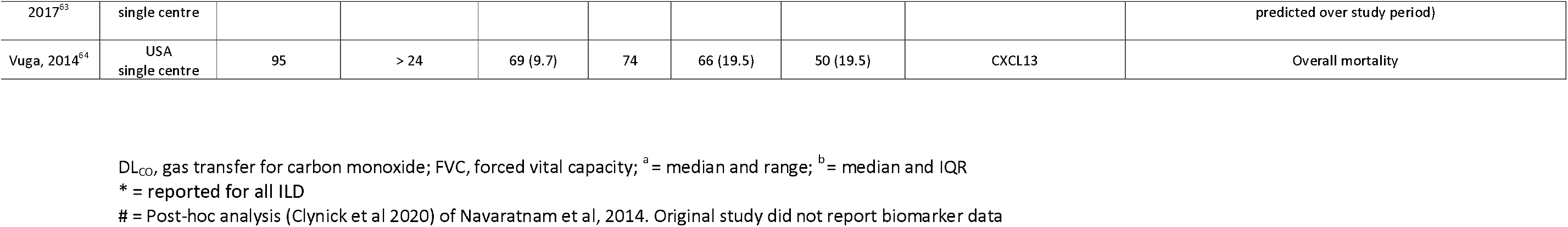
Methodological characteristics of included studies with baseline participant characteristics and outcome data. Age, baseline FVC and baseline DLCO reported as mean (standard deviation) unless otherwise stated.

**Figure 1.**
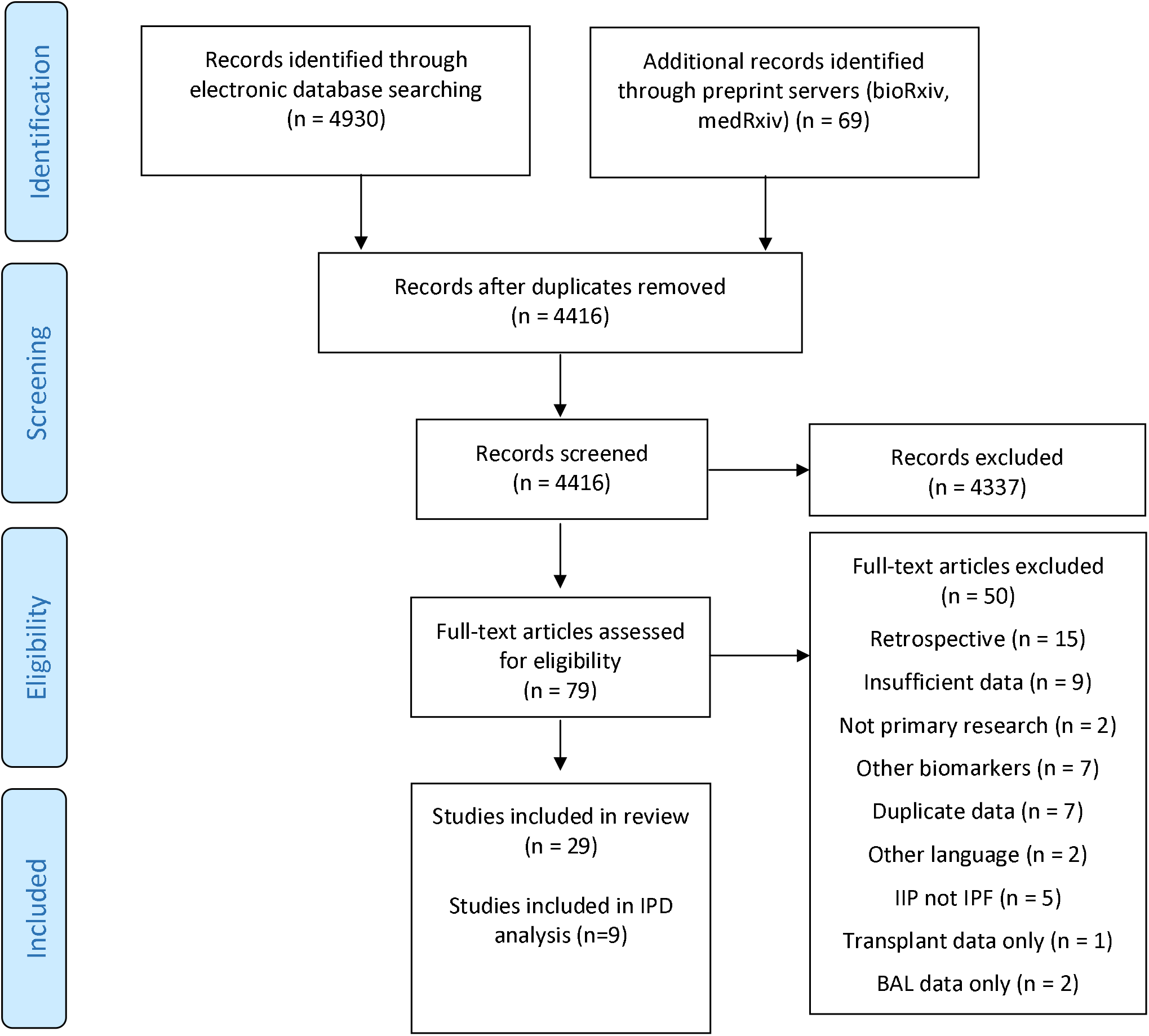
Flow diagram illustrates systematic search and screening strategy, including numbers of studies meeting eligibility criteria and numbers excluded.

Risk of bias assessment of the retrieved studies identified limitations and a number of possible biases (Figure 2, Table S1). Most studies defined the study population specifically with clear inclusion/exclusion criteria. Biomarkers were measured consistently using the same sample matrices (plasma or serum) across included participants in each study, although details of assay platforms used to measure the analytes were frequently unreported. Outcome data were measured objectively and applied consistently to all study participants. In approximately half of the studies, possible confounders were not measured, and findings were often reported using data-dependent cut off thresholds. For studies included in the MMP-7 meta-analysis, publication bias was not observed (Figure S3 and S4).

**Figure 2.**
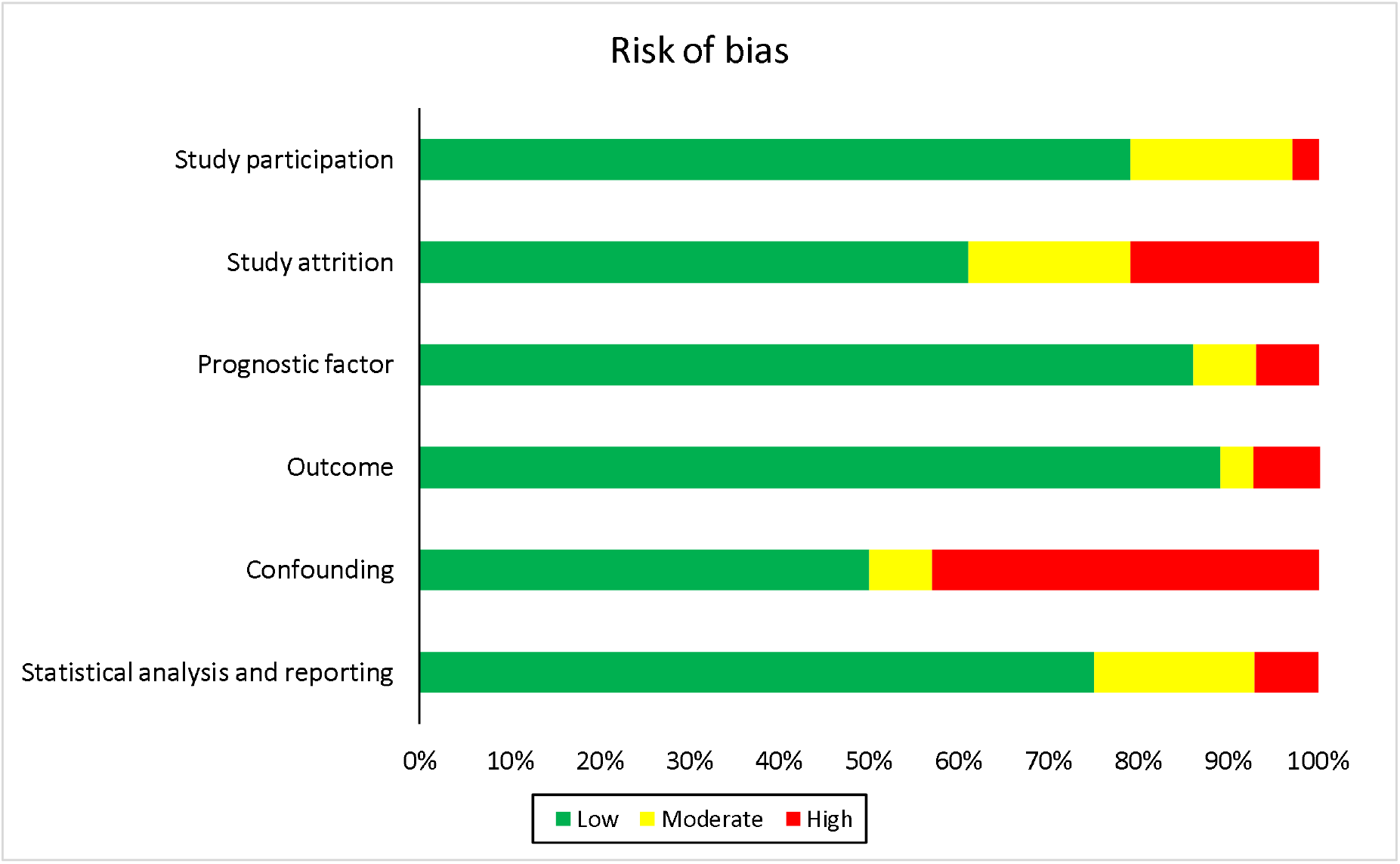
Risk of bias assessment for included studies. The risk of bias across studies was rated as low, moderate or high risk in six categories using the QUIPs tool.

## Association between blood biomarkers and clinical outcomes

### Baseline blood biomarkers that predict mortality

The primary outcome of mortality was evaluated in 22 studies, reporting outcomes from 3284 participants, with sample sizes ranging from 18-438 and follow up periods from 8-60 months. Ten studies evaluated the relationship between mortality and MMP-7, with IPD available for eight studies totalling 1492 participants. Meta-analysis demonstrated elevated baseline MMP-7 values were associated with a 23% increased risk of overall mortality [adjusted HR (aHR) 1.23 per standard deviation (SD) increase, 95%CI 1.03;1.48, I^2^=64.3%) (Figure 3A), though there was substantial statistical heterogeneity which could not be explained by variability in the factors assessed (Table S2). When mortality at 12 months was examined specifically, baseline MMP-7 levels were inconclusively associated with death (aHR 1.33 per SD increase, 95%CI 0.99;1.78, I^2^=59.6%) (Figure 3B). Applying the GRADE framework (Table S8), we rate the confidence in mortality estimates with moderate certainty (Table S8). Where IPD was unavailable, MMP-7 values above 5.7ng/mL were associated with increased mortality (aHR 2.18 95%CI 1.1;4.32) over a median follow up of 19 months in a study of 438 participants.^22^ A further study of 57 participants found MMP-7 levels did not predict death^23^ (Table S3).

**Figure 3.**
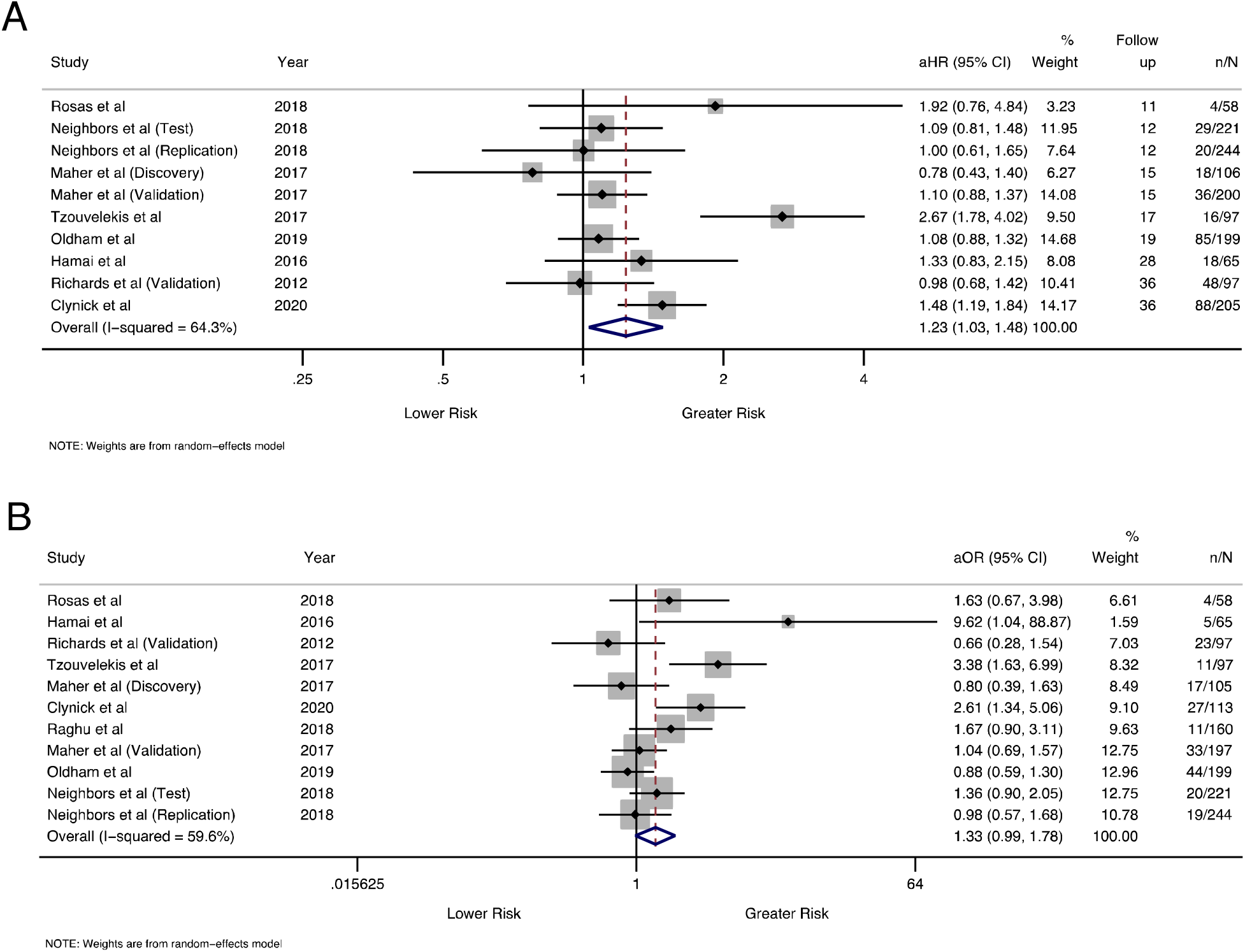
Mortality forest plot. A – Overall mortality. B: Mortality at 12 months. Effect sizes with 95% confidence intervals per standard deviation increase in baseline MMP-7. Study follow up time shown in months. n denotes the number of deaths, and N represents the total number of participants included per study.

Whilst many of the remaining 14 biomarkers showed an association with mortality, findings were inconsistent and inconclusive (Figure 6 and S3). Study follow up times were inconsistent, effect sizes varied with wide confidence intervals, and estimates were often unadjusted for important covariates.

### Change in biomarkers predicting mortality

Three studies totalling 498 participants explored the association between MMP-7 change over three months and mortality.^24 25^ IPD meta-analysis showed no association with mortality (aHR 1.00, 95%CI 0.99;1.02, I^2^=53.3%), nor when mortality was censored at 12 months (aOR 1.00, 95%CI 0.99;1.01, I^2^=37.4%) (Figures S5 and S6).

Three publications reporting from the same cohort evaluated the relationship between longitudinal biomarker measurement and mortality.^10 26 27^ In both discovery and validation cohorts, a rise in CA-125 over three-months doubled the risk of death, but the remaining biomarkers were not predictive of mortality (Figure 6 and S4). A validation cohort of 145 participants demonstrated replication of rising neoepitopes degraded by matrix metalloproteinases (C1M, C3M, C6M and CRPM), but the rate of change of collagen synthesis peptides was not associated with mortality.^27^

### Baseline biomarkers that predict disease progression and change in FVC

Disease progression in IPF was reported in 19 studies and 13 studies evaluated the role of serum biomarkers in predicting FVC change alone. Ten studies measured MMP-7 levels as markers of disease progression, with eight studies totalling 1383 participants included in the IPD meta-analysis. Disease progression was standardised as greater than or equal to 10% relative decline in FVC or death within 12 months of baseline. Meta-analysis demonstrated baseline MMP-7 was associated with disease progression (aOR 1.27 per SD increase, 95%CI 1.11;1.46, I^2^=5.9%) (Figure 4). Whilst heterogeneity was low, meta-regression identified sample assay techniques (ELISA vs. other) to be a source of heterogeneity. In subgroup analysis according to assay, the odds ratio for disease progression was estimated at 1.56 per SD increase (95%CI 1.26;1.82, I^2^=0%) when restricted to studies using ELISA (Figure S7). When the relationship between baseline MMP-7 and relative change in FVC at 12 months was examined specifically in six studies of 891 participants, meta-analysis indicated that a 1 standard deviation greater baseline MMP-7 was associated with a −0.85% relative change in 12-month FVC percent predicted (95%CI −1.65; −0.05, I^2^=0%) (Figure 5). We assessed our findings for disease progression and change in FVC outcomes with high certainty (Table S8). For studies not included in IPD meta-analysis, baseline MMP-7 values above 3.8ng/mL doubled the risk of disease progression (aHR 2.2 95%CI 1.4;3.7) over a median follow-up of 19 months in 211 participants.^28^ In a further study of 57 participants, MMP-7 did not predict disease progression (Table S6).

**Figure 4.**
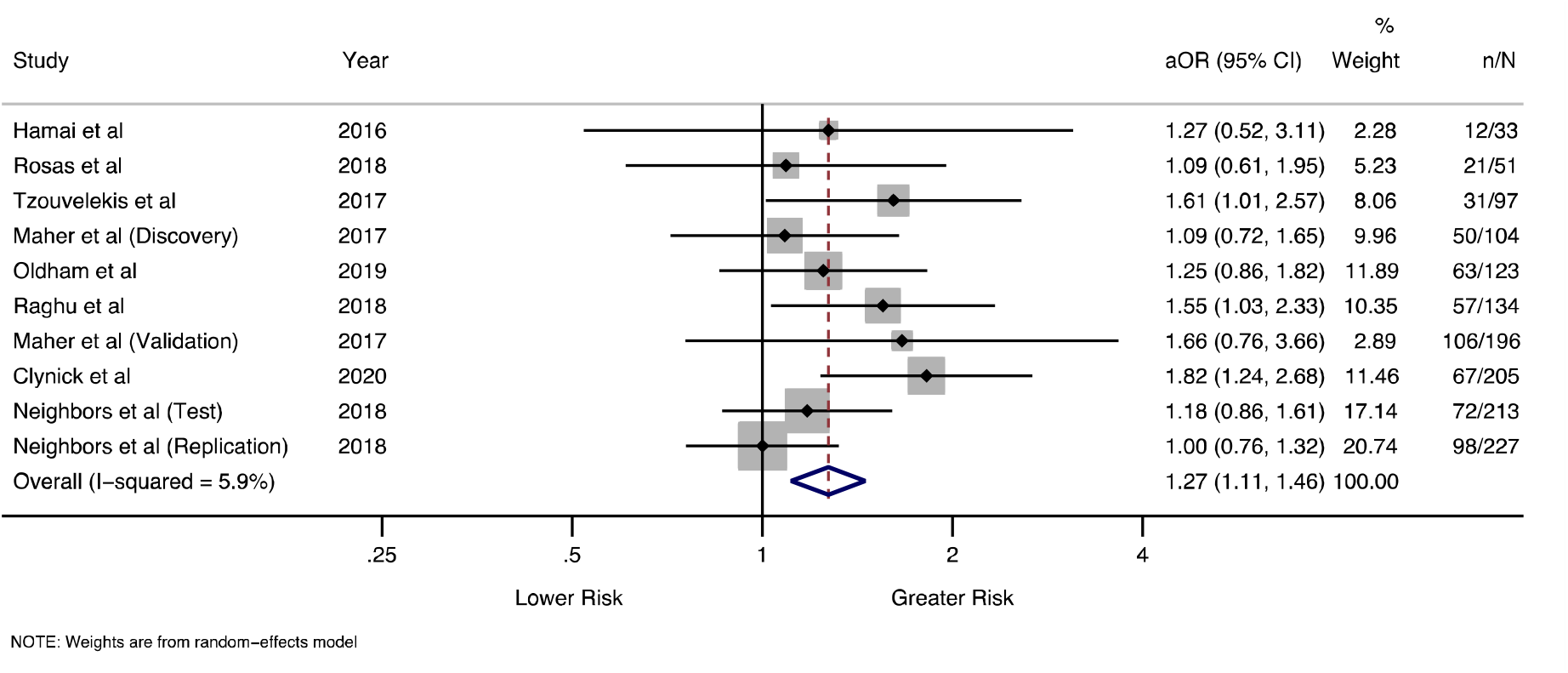
Disease progression forest plot. Pooled odds ratios with 95% confidence intervals for risk of disease progression, per standard deviation increase in baseline MMP-7. n denotes the number of progressors, and N represents the total number of participants included in the analysis per study.

**Figure 5.**
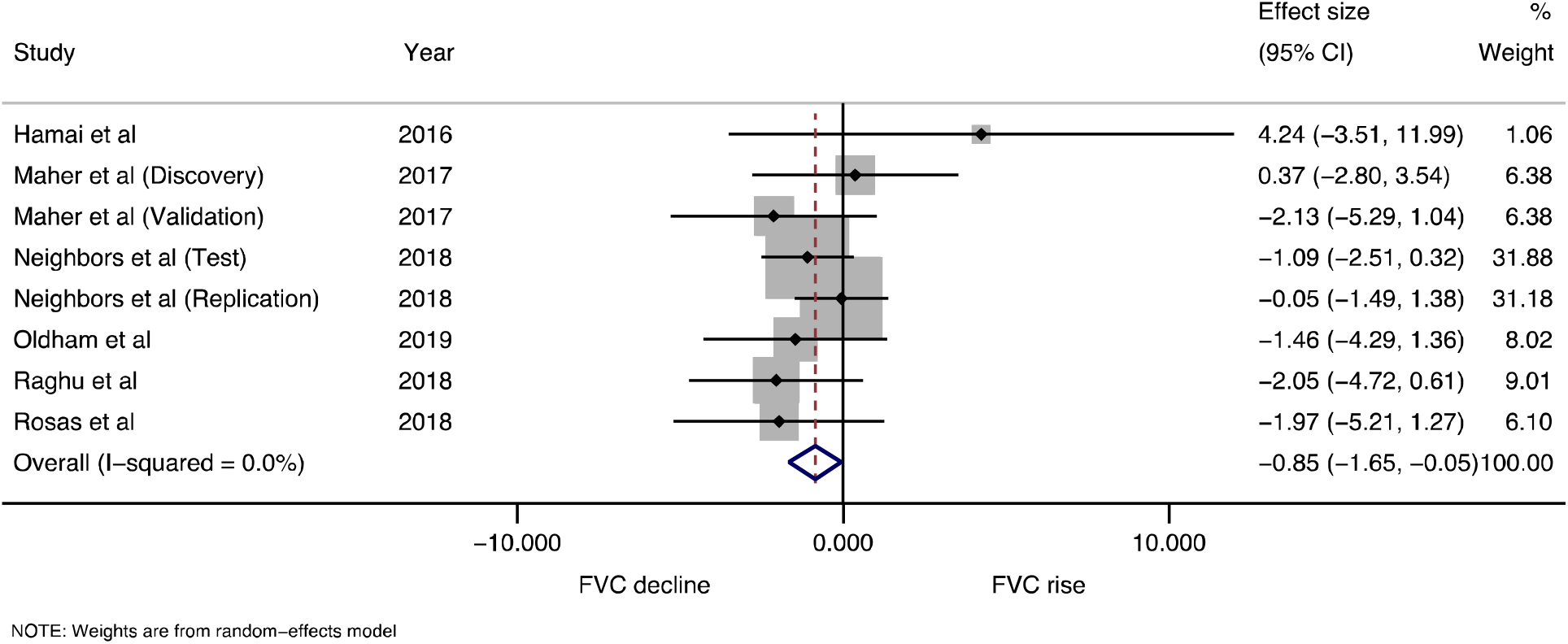
Relative change in FVC% percent predicted forest plot Pooled effect size with 95% confidence intervals for FVC% percent predicted relative change at 12 months, per standard deviation increase in baseline MMP-7.

**Figure 6.**
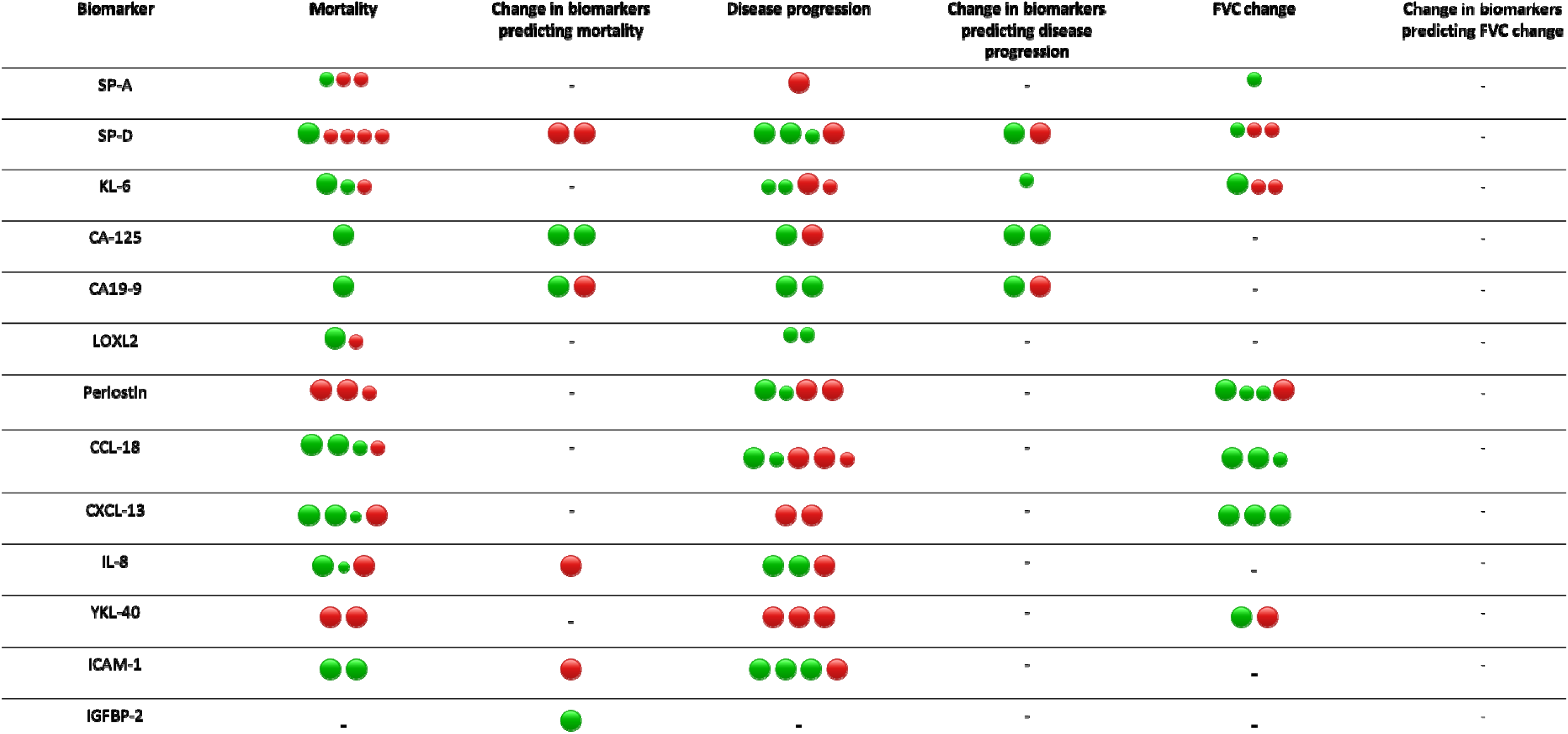
Summary of study results. Each dot represents a study (or individual cohort in studies with more than one cohort). Green dots represent studies showing an association between the biomarker and outcome, and red dots represent studies where no association was found. Larger circles represent studies with a sample size > 100 participants, and smaller circles represent studies with sample sizes smaller than 100 participants. Outcomes where no studies were found for the listed biomarker are represented with a dash (-).

In studies of other biomarkers, none were consistently predictive of disease progression. However, there was significant heterogeneity in adopted definitions of disease progression, with lung function indices, mortality, transplant and acute exacerbations included in various combinations at non-unified time points (Figure 6 and S5).

### Change in biomarkers predicting disease progression

Three studies totalling 481 participants investigating the association between MMP-7 change over three months and disease progression were included in IPD meta-analysis. Change in MMP-7 over three-months was not associated with disease progression (aOR 1.00 per percent increase, 95%CI 0.99;1.01, I^2^=22.5%) (Figure S8), nor with change in FVC over 12 months (effect size 0.01% increase per percent MMP-7 increase 95%CI −0.07;0.08, I^2^=60.8%) (Figure S9). In a study of 211 participants that could not be included in IPD meta-analysis, a two-fold change in MMP-7 over four months was associated with doubling the risk of disease progression.^28^

In one study, participants with progressive disease had rising concentrations of CA-125 over 3 months compared to those with stable disease, but no relationship was replicated for other biomarkers.^10^ (Figure 6, S6 and S7)

## DISCUSSION

This systematic review of prospective studies summarises the association between MMP-7 and clinical outcomes in patients with untreated IPF, as well as estimates reported with other blood biomarkers. Our results of IPD meta-analysis demonstrate baseline MMP-7 levels predicted all-cause mortality and disease progression and correlated with FVC percent predicted change over 12 months. MMP-7 levels did not seem to change longitudinally over three months, with no association observed with any of the measured outcomes. However, a study not included in quantitative synthesis suggested that in those individuals where MMP-7 does rise, there may be an associated risk in progression ^28^. We assess the certainty of our findings using GRADE (Table S8) and rate mortality outcomes with moderate certainty and disease progression and change in FVC outcomes with high certainty.

The current paradigm for the pathogenesis of IPF suggests a complex interplay of a dysfunctional alveolar epithelium with aberrant wound healing leading to chronic fibro-proliferation following repeated epithelial micro-injury.^29^ Dysfunctional epithelial cells contribute to fibrogenesis by secreting profibrotic mediators including matrix-metalloproteinases (MMPs).^30^ MMPs are zinc-containing endopeptidases responsible for degrading multiple components of extracellular matrix, activating biological mediators, and facilitating cell migration.^31^ Overexpression of MMP-7 would therefore be consistent with increased disease activity and fibrogenesis, supporting its role as a potential prognostic biomarker in IPF.

Due to heterogeneity in study designs and reported outcomes, there was insufficient data for quantitative analysis in non-MMP-7 studies. Whilst many biomarkers showed an association with mortality in single studies, replication of effects across studies was weak. Short-term changes in biomarker concentrations over three-months were often not associated with specified clinical outcomes suggesting further studies are needed before such biomarkers can be adopted clinically.

We highlight sources of considerable bias and variability between blood biomarker studies in IPF. The lack of sample size calculations in many studies suggests power was not defined in prespecified designs. Studies were typically observational and cohort in nature, of relatively modest size, but others included participants from placebo arms in interventional randomised clinical trials. Interventional trials had differing participant exclusion criteria according to study objectives, which may limit generalisability. A number of different laboratory techniques were applied to measure biomarker levels across studies, with very few studies reporting detailed assay information, particularly with regards to measures of precision, and there was inconsistency in thresholds defining positive and negative biomarker result. Analysis of heterogeneity in IPD meta-analysis indicated that assay type was a significant contributor to heterogeneity, particularly in estimates of disease progression.

In view of inter-study variability and inconsistency in the reporting of outcomes and effect measures, we sought to address some of these limitations by presenting our findings narratively and obtaining IPD to enable meta-analysis of MMP-7 as a prognostic factor, a particular strength of this review. This method enabled analysis of biomarkers as continuous variables transformed to z-scores to overcome assay variability, supported standardised definition of outcomes, and consistent adjustment for important covariates, which improves the reliability of our findings. We performed two-stage IPD meta-analysis, which does not assess study estimate and effects simultaneously although is considered to produce unbiased estimates,^32^ and enabled modelling IPD from 1492 participants across separate secure servers and portals.

There are limitations to this review. Whilst language restrictions were not applied, two articles in Japanese were excluded as they could not be translated to English to assess inclusion criteria. We included only those studies where participants were diagnosed according to international consensus guidelines, supporting the robustness and generalisability of our findings to IPF patients. We excluded studies in IIPs not specific to IPF, which limits generalisability to non-IPF ILDs, although ongoing studies exploring shared mechanistic pathways will provide further insight.^33^ Furthermore, by focussing on untreated IPF patients as molecular consequences of treatment may be unknown, our results do not address theranostic value in relation to anti-fibrotics. There was significant statistical heterogeneity in some of the outcomes, and therefore these should be interpreted with caution. We were unable to explain all the residual heterogeneity using the factors we assessed. IPD was not obtained from a limited number of suitable studies, and we report these findings narratively.

In summary, we demonstrate through IPD meta-analysis that baseline MMP-7 levels predicted overall mortality and disease progression in patients with untreated IPF. However, short term changes in MMP-7 over three-months offered limited prognostic value in the absence of an empirical threshold. Further research should focus on exploring the relationship between IPF pharmacotherapy and MMP-7, particularly to identify whether changes in MMP-7 levels may present a biomarker of therapeutic response. From a clinical perspective, MMP-7 should be considered for implementation as a prognostic tool at the point of diagnosis, especially where lung function testing is cumbersome or unavailable. Whilst a number of other blood biomarkers have been studied for predicting prognosis, there is currently insufficient replication to enable adoption into clinical testing. Further biomarker research should focus on rigorously designed longitudinal studies with discovery and validation cohorts, using validated biomarker assays and standardised endpoints.

## Data Availability

Meta-analysis of published data

## Acknowledgements

This publication is based on research using data from data contributors Boehringer Ingelheim and Genentech Inc. that has been made available through Vivli, Inc. Vivli has not contributed to or approved, and is not in any way responsible for, the contents of this publication. Boehringer Ingelheim was given the opportunity to review the abstract for medical and scientific accuracy, as well as intellectual property considerations.

This publication also uses data from Sanofi made available through ClinicalStudyDataRequest.com. The interpretation and reporting of research using these data are solely the responsibility of the authors and do not necessarily represent the official views of ClinicalStudyDataRequest.com or Sanofi.

The authors would also like to thank the following individuals for their invaluable support in providing access to individual patient data: Dr Hiroshi Ivamoto (Hiroshima University, Japan), Professor Naftali Kaminski (Yale School of Medicine, USA), Dr Margaret Neighbors (Genentech, Inc., USA), Dr Justin Oldham (University of California, USA), Professor Ganesh Raghu (University of Washington Medical Centre USA), Dr Ivan Rosas (Brigham and Women’s hospital, USA), Dr Argyrios Tzouvelekis (Alexander Fleming Biomedical Sciences Research Center, Greece), Dr Y Zhang (University of Pittsburgh, USA),

