## Supplementary File for "An individual participant data meta-analysis of prognostic blood biomarkers in IPF"

**Supplementary Material**

Figure 1 - MEDLINE search strategy

Figure 2 – Letter to authors for individual participant data

Figure 3 – Funnel plots for baseline MMP-7

Figure 4 – Funnel plots for change in MMP-7 over 3 months

Figure 5 – Forest plot for change in MMP-7 over 3 months and overall mortality

Figure 6 – Forest plot for change in MMP-7 over 3 months and 12 months mortality

Figure 7 – Forest plot for baseline MMP-7 and disease progression separated by ELISA and non-ELISA

Figure 8 – Forest plot for change in MMP-7 over 3 months and disease progression

Figure 9 – Forest plot for change in MMP-7 over 3 months and relative change in FVC at 12 months

Table 1 – Risk of bias for included studies per individual study

Table 2 – Meta-regression for variables assessed

Table 3 – Table of mortality outcomes for baseline biomarkers

Table 4 – Table of mortality outcomes for short term change in biomarkers

Table 5 – Table of disease progression definition and outcomes for baseline biomarkers

Table 6 – Table of disease progression definitions and outcomes for short term change in biomarkers

Table 7 – Table of FVC change definitions and outcomes for baseline biomarkers

Table 8 – GRADE rating

### Search strategy: MEDLINE

1. idiopathic pulmonary fibros*.mp.

2. pulmonary fibros*.mp.

3. Pulmonary Fibrosis/ or Idiopathic Pulmonary Fibrosis/

4. cryptogenic fibrosing alveolitis.mp.

5. usual interstitial pneumonia*.mp.

6. Fibrosing alveolitis.mp.

7. Idiopathic Interstitial Pneumonia*.mp.

8. Interstitial pneumonia*.mp.

9. Idiopathic interstitial lung disease.mp.

10. Chronic interstitial pneumonia*.mp.

11. 1 or 2 or 3 or 4 or 5 or 6 or 7 or 8 or 9 or 10

12. Mucin-1/

13. KL-6.mp.

14. krebs von den lungen-6.mp.

15. SP-A.mp.

16. Pulmonary Surfactant-Associated Protein A/

17. Pulmonary Surfactant-Associated Protein D/

18. Pulmonary Surfactants/

19. SP-D.mp.

20. surfactant protein*.mp.

21. CA-125 Antigen/ or CA125.mp.

22. cancer antigen 125.mp.

23. mucin 16.mp.

24. CA-19-9 Antigen/ or CA19-9.mp.

25. cancer antigen 19-9.mp.

26. carbohydrate antigen 19-9.mp.

27. Matrix Metalloproteinase 1/ or MMP-1.mp.

28. Matrix Metalloproteinase 7/ or MMP-7.mp.

29. matrix metalloproteinase.mp. or Matrix Metalloproteinases/

30. LOXL2.mp.

31. Protein-Lysine 6-Oxidase/

32. protein-lysine 6-oxidase.mp.

33. periostin.mp.

34. Osteoblast-specific factor 2.mp.

35. Epitopes/ or Neoepitope*.mp.

36. Chemokines, CC/ or CCL18.mp.

37. Chemokine CCL18.mp.

38. Chemokines, CC/ or CC-chemokine ligand 18.mp.

39. IL-8.mp. or Interleukin-8/

40. Interleukin-8.mp.

41. CXCL8.mp.

42. Chemokine ligand 8.mp.

43. Chitinase-3-Like Protein 1/ or YKL-40.mp.

44. CHI3L1.mp.

45. Chitinase-3-Like Protein 1/ or Chitinase-3-like protein 1.mp.

46. IGFBP-2.mp. or Insulin-Like Growth Factor Binding Protein 2/

47. Insulin like growth factor binding protein 2.mp.

48. ICAM-1.mp. or Intercellular Adhesion Molecule-1/

49. VEGF.mp. or Vascular Endothelial Growth Factor A/

50. HSP70 HEAT-SHOCK PROTEINS/ or HSP70.mp.

51. LEPTIN/ or Leptin.mp.

52. CXCL13.mp. [mp=title, abstract, original title, name of substance word, subject heading word, floating sub-heading word, keyword heading word, protocol supplementary concept word, rare disease supplementary concept word, unique identifier, synonyms]

53. Chemokine CXCL13/ or C-X-C motif chemokine 13.mp.

54. Forced Vital Capacity.mp. or Vital Capacity/

55. FVC.mp.

56. Forced Expiratory Volume/ or FEV1.mp.

57. forced expiratory volume.mp.

58. 6 minute walk.mp.

59. Six minute walk.mp.

60. Walk Test/

61. walk test.mp.

62. 6MWT.mp.

63. 6MWD.mp.

64. Pulmonary diffusing capacity.mp. or Pulmonary Diffusing Capacity/

65. Diffusion capacity for carbon monoxide.mp.

66. DLCO.mp.

67. Transfer factor.mp. or Transfer Factor/

68. Gas transfer.mp.

69. TLCO.mp.

70. KCO.mp.

71. PHYSIOLOGY/

72. Physiolog*.mp.

73. SPIROMETRY/

74. spiromet*.mp.

75. biomarkers.mp. or BIOMARKERS/

76. ((Serum or clinical or immun* or lab or laboratory or biochemical or biological) and marker*).mp.

77. or/12-76

78. prognosis.sh.

79. diagnosed.tw.

80. cohort:.mp.

81. predictor:.tw.

82. death.tw.

83. exp models, statistical/

84. disease progression.sh.

85. disease progression.mp.

86. 78 or 79 or 80 or 81 or 82 or 83 or 84 or 85

87. 11 and 77 and 86

88. limit 87 to humans

Supplementary Figure 1 – MEDLINE search strategy (last search carried out on 12th November 2020)

Copy of email sent to authors

We would be very grateful for your assistance in undertaking a robust meta-analysis. The team at University of Nottingham (UK), led by Prof Gisli Jenkins, are conducting a systematic review and meta-analysis of blood biomarkers in IPF. The protocol for the study can be found on PROSPERO: https://www.crd.york.ac.uk/prospero/display_record.php?RecordID=120402

As part of the review, we will conduct a meta-analysis of the association of MMP-7 levels with mortality in IPF. We have chosen this biomarker because there is sufficient published data to make it feasible and useful.

To assist with this, we would be extremely grateful if you could kindly provide us with individual patient data from your highly relevant study entitled "…" published in …

We also note significant heterogeneity in disease progression definitions across individual studies, and therefore hope to meta-analyse MMP-7 level associations with a shared definition based on FVC and mortality and would also appreciate data to assist with this. We appreciate the inconvenience such requests entail, and we would like to make the process as smooth as possible, we will of course acknowledge all support.

The attached excel spreadsheet highlights the anonymised data we are seeking for each patient, where available:

- MMP-7 level (baseline and 3-months)
- Assay method (type of assay used)
- Sample type (serum or plasma)
- Age
- Gender (M or F)
- Follow up time (days)
- Dead or alive at end
- Time to death (days)
- Baseline FVC (% predicted)
- 3-month FVC (% predicted)
- 12-month FVC (% predicted)
- Smoking (ever or never)

Thank you for your help, we look forward to communicating with you further.

Supplementary Figure 2 – copy of message sent to authors for individual participant data. A minimum of three reminders, 4 weeks apart were sent.

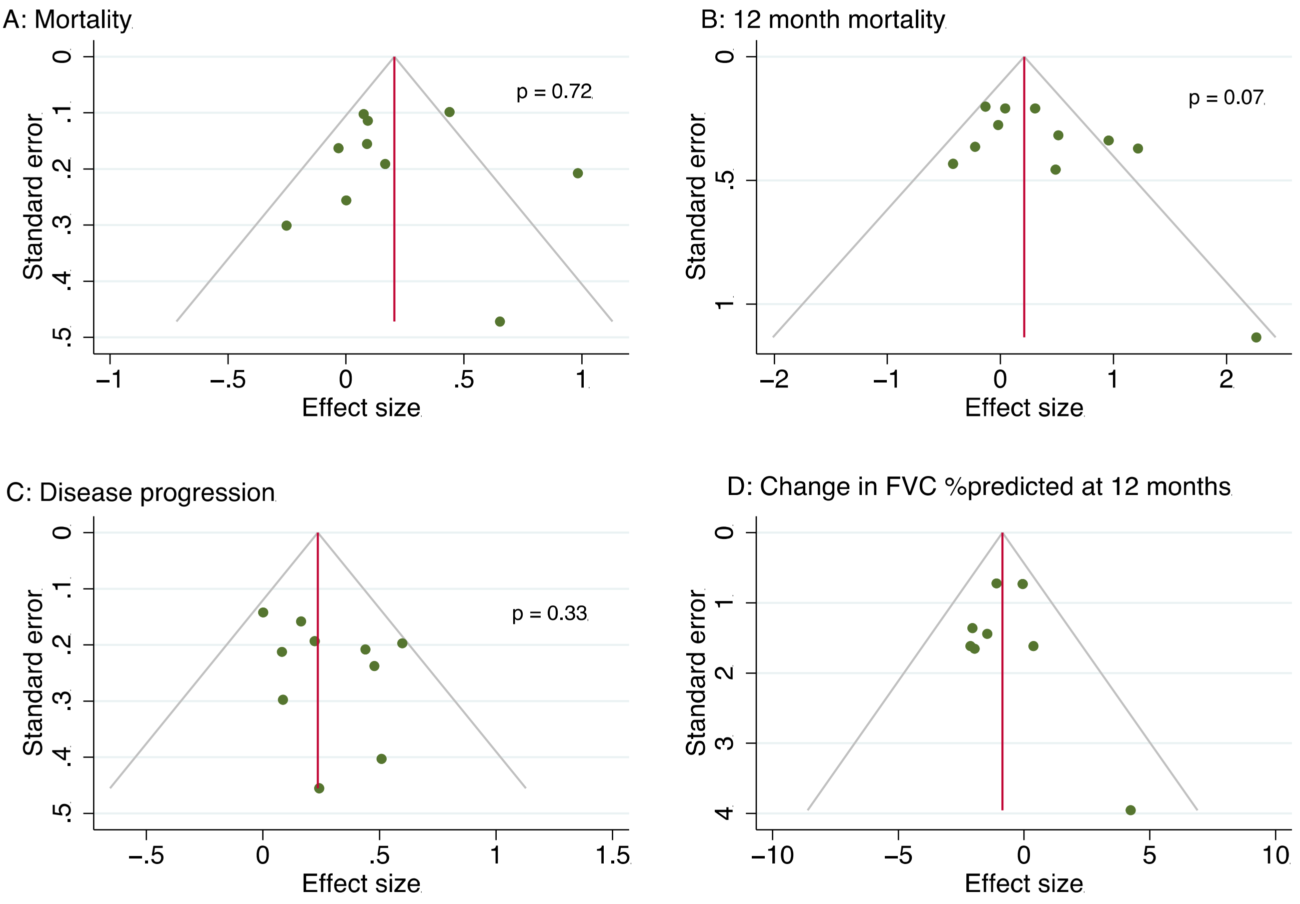

Supplementary Figure 3 – Funnel plots for outcomes evaluated in baseline MMP-7 IPD meta-analysis. A: overall mortality, B: 12-month mortality, C: Disease progression, D: Change in percent predicted FVC at 12 months. Publication bias assessed using Egger's test for outcomes with at least ten studies, and p values presented next to funnel plot.

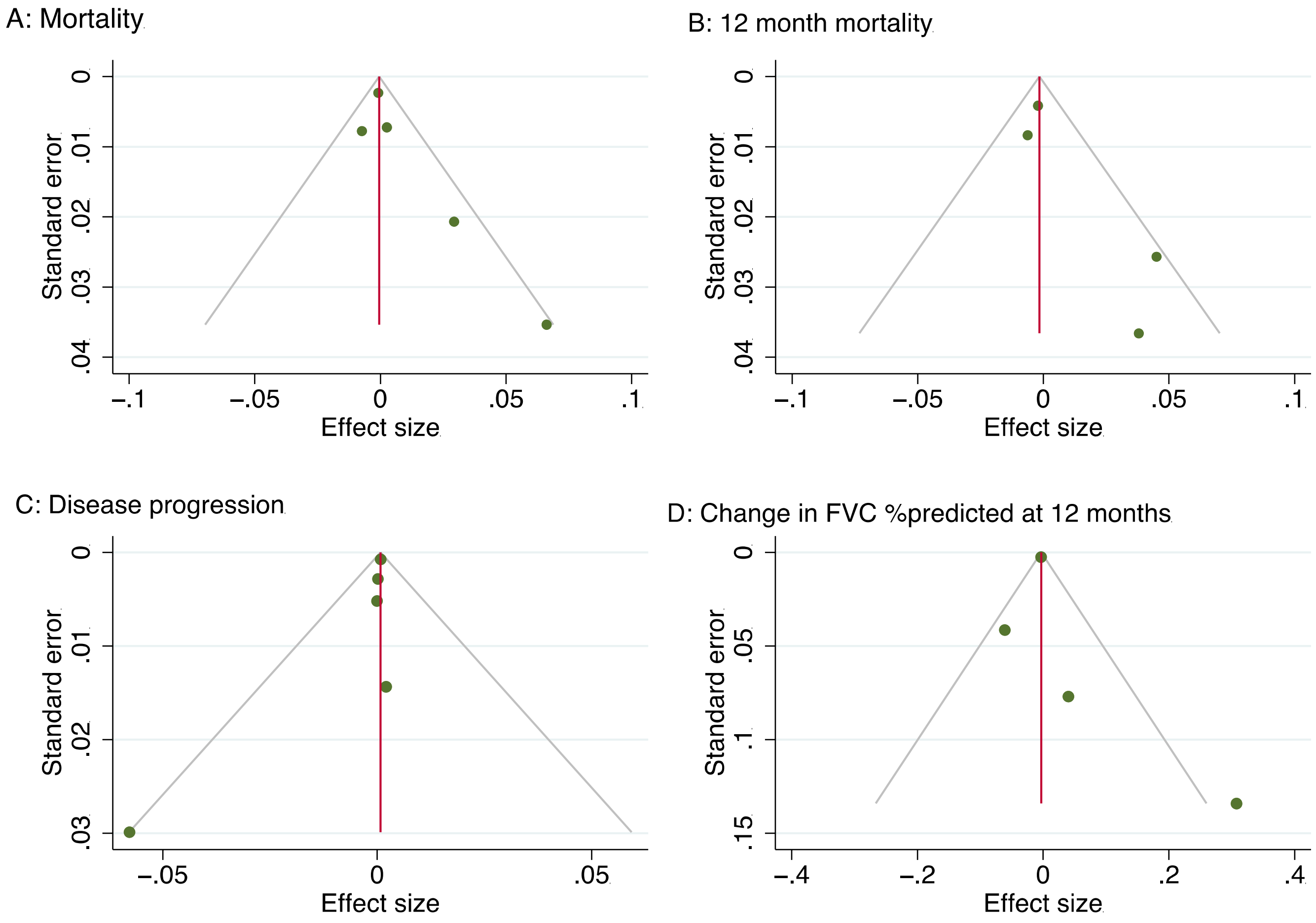

Supplementary Figure 4 – Funnel plots for outcomes evaluated for three-month change in MMP-7 IPD meta-analysis. A: overall mortality, B: 12-month mortality, C: Disease progression, D: Change in percent predicted FVC at 12 months.

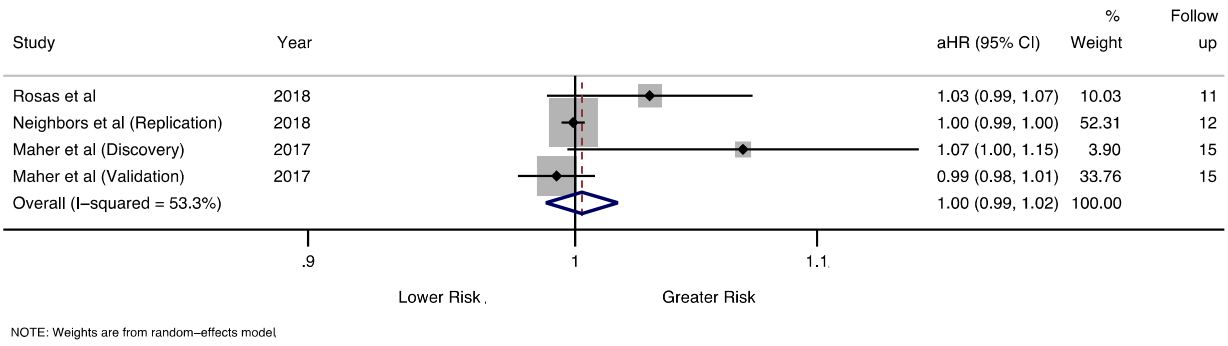

Supplementary Figure 5 - Pooled hazard ratios with 95% confidence intervals for risk of overall mortality, per percent relative increase in MMP-7 from baseline to three months. Study follow up time shown in months. n denotes the number of deaths, and N represents the total number of participants included per study.

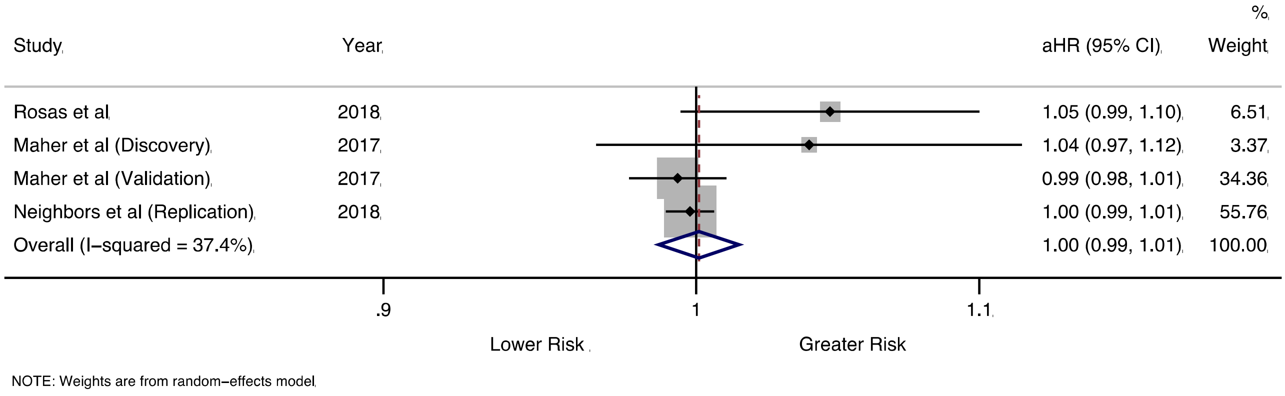

Supplementary Figure 6 - Pooled hazard ratios with 95% confidence intervals for risk of mortality at 12 months, per percent relative increase in MMP-7 from baseline to three months. n denotes the number of deaths, and N represents the total number of participants included per study.

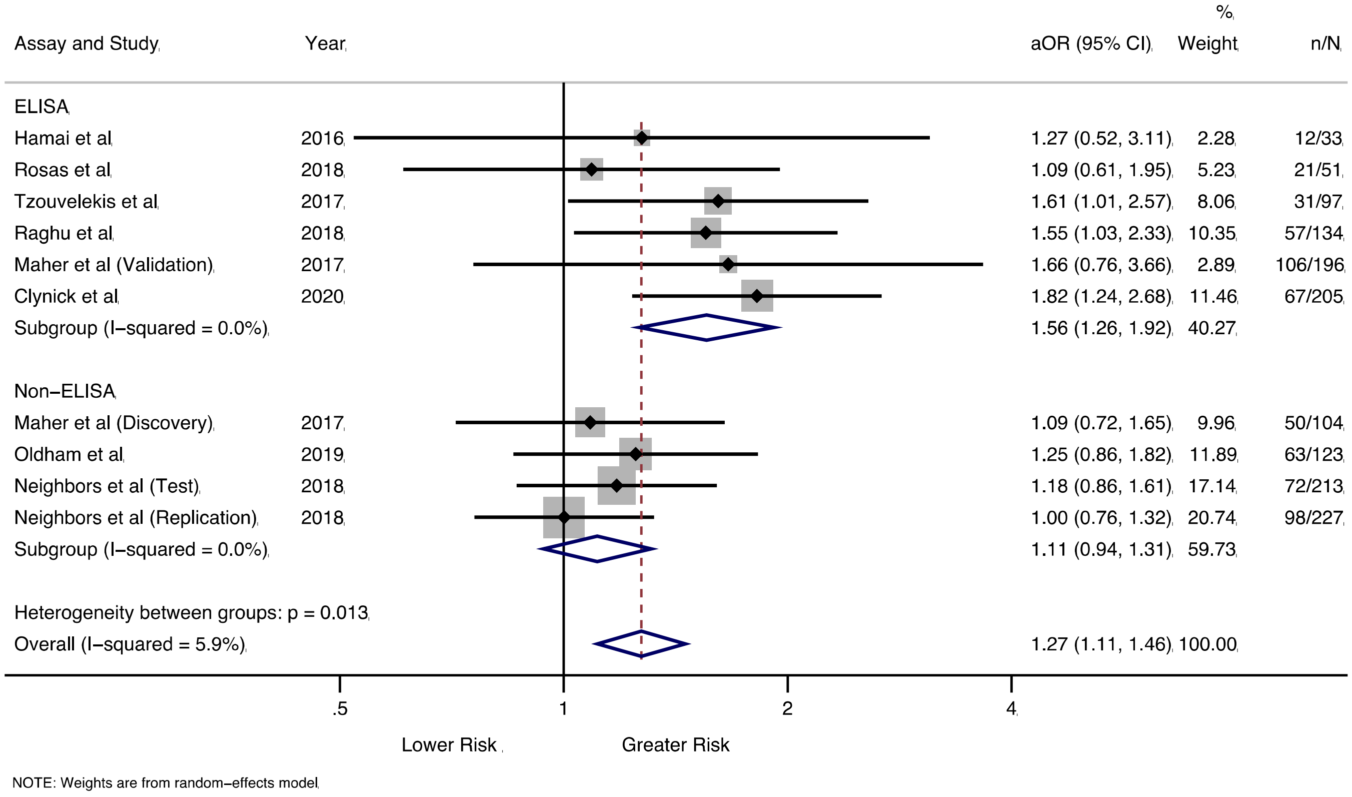

Supplementary Figure 7 – Pooled odds ratios with 95% confidence intervals for risk of disease progression, per standard deviation increase in baseline MMP-7. Separated by ELISA and non-ELISA measurements. n denotes the number of progressors, and N represents the total number of participants included in the analysis per study.

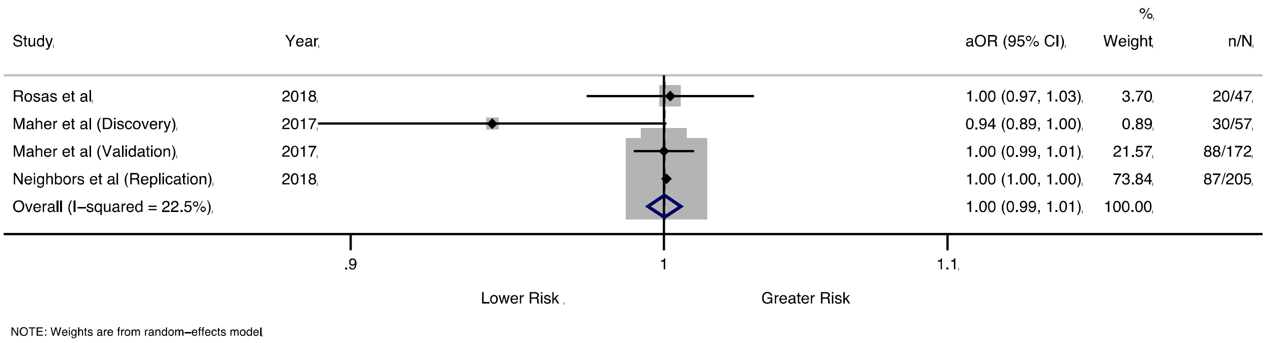

Supplementary Figure 8 – Pooled odds ratios with 95% confidence intervals for risk of disease progression, per percent relative increase in baseline MMP-7 to three months. n denotes the number of progressors, and N represents the total number of participants included in the analysis per study.

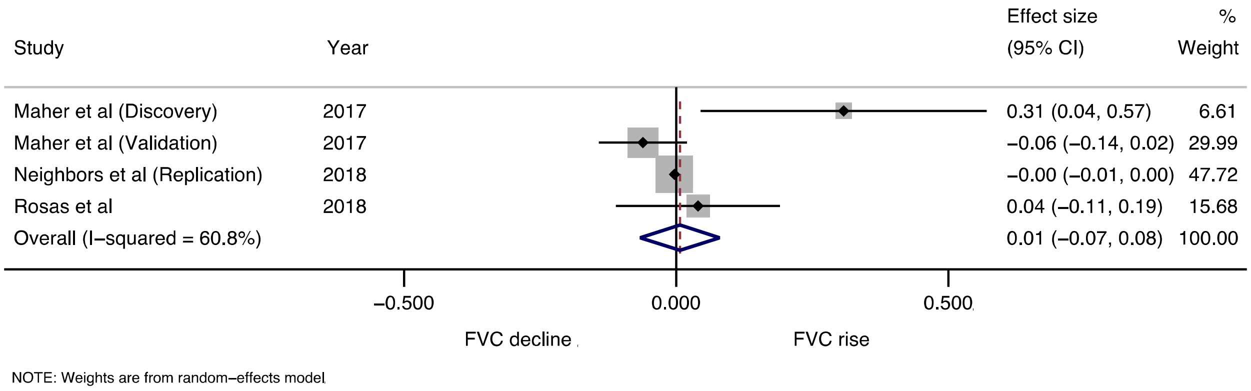

Supplementary Figure 9 – Pooled effect size with 95% confidence intervals for relative change in FVC at 12 months, per percent relative increase in baseline MMP-7 to three months.

| Study | Study participation | Study attrition | Prognostic factor | Outcome | Confounding | Statistical analysis and reporting |
|---|---|---|---|---|---|---|
| Bauer, 2017 | Low | Low | Moderate | Low | High | Low |
| Chien, 2014 | Low | Low | Low | Low | Moderate | Low |
| Collard, 2010 | Low | Low | Low | Low | High | Low |
| Doubkova, 2016 | Moderate | High | High | High | High | High |
| Gui, 2020 | Low | Low | Low | Moderate | High | Low |
| Hamai, 2016 | Moderate | Moderate | Low | Low | Low | Low |
| Hoyer, 2020 | High | High | High | Low | High | High |
| Jiang, 2018 | Low | Low | Low | Low | High | Low |
| Jenkins, 2015 | Low | Moderate | Low | Low | Low | Low |
| Kennedy, 2015 | Moderate | Low | Low | Low | High | Moderate |
| Kinder, 2009 | Low | Low | Low | Low | Low | Low |
| Maher, 2017 | Low | Moderate | Low | Low | Low | Low |
| Naik, 2012 | Low | Low | Low | Low | Low | Low |
| Navaratnam, 2014/Clynick, 2020 | Low | Moderate | Low | Low | Low | Low |
| Neighbors, 2018 | Low | Low | Low | Low | Low | Low |
| Ohshimo, 2014 | Low | Low | Low | Low | Low | Low |
| Ohta, 2017 | Low | High | Low | Low | High | Low |
| Okamoto, 2011 | Low | High | Low | Low | Low | Moderate |
| Oldham, 2019 | Low | High | High | Low | High | Moderate |
| Organ, 2019 | Low | Moderate | Low | Low | Low | Low |
| Papiris, 2018 | Low | Low | Low | Low | High | Moderate |
| Peljto, 2013 | Low | Low | Moderate | Low | Low | Low |
| Prasse, 2009 | Moderate | Low | Low | Low | Low | Low |
| Richards, 2012 | Low | Low | Low | Low | Moderate | Low |
| Rosas, 2018 | Low | Low | Low | Low | High | Moderate |
| Sokai, 2015 | Low | Low | Low | Low | High | Low |
| Tzouvelekis, 2017 | Low | Low | Low | Low | Low | Low |
| Vuga, 2014 | Moderate | High | Low | High | Low | Low |

Supplementary Table 1 – Risk of bias assessment for included studies. The risk of bias across studies was rated as low, moderate or high risk in six categories using the QUIPs tool.

| Baseline MMP-7 | | | | | | | | |
|---|---|---|---|---|---|---|---|---|
| Variables | Overall mortality (n=1492) | | 12-month mortality (n= 1492) | | Disease progression (n= 1383) | | Change in FVC percent predicted over 12 months (n=891) | |
| | $R^2$ (%) | P value | $R^2$ (%) | P value | $R^2$ (%) | P value | $R^2$ (%) | P value |
| Design (cohort vs. RCT) | 0.00 | 0.747 | 0.00 | 0.388 | 0.00 | 0.159 | 0.00 | 0.988 |
| Assay (ELISA vs. other) | 18.45 | 0.088 | 25.4 | 0.075 | 100 | 0.013 | 0.00 | 0.235 |
| Sample (Serum vs. plasma) | 0.00 | 0.98 | 0.00 | 0.483 | 71.35 | 0.1875 | 0.00 | 0.502 |
| IPF consensus (2011 vs. other) | 0.00 | 0.983 | 0.00 | 0.87 | 100 | 0.05 | N/A | N/A |
| Centre (single vs. multi) | 9.05 | 0.1995 | 0.00 | 0.293 | 6.23 | 0.418 | 91.14 | 0.195 |
| Publication type (peer reviewed) | 0.00 | 0.922 | 0.00 | 0.893 | 47.51 | 0.212 | 0.00 | 0.659 |
| **Change in MMP-7 over 3 months** | | | | | | | | |
| Variables | Overall mortality (n=498) | | 12-month mortality (n=498) | | Disease progression (n= 481) | | Change in FVC percent predicted over 12 months (n= 481) | |
| | $R^2$ (%) | P value | $R^2$ (%) | P value | $R^2$ (%) | P value | $R^2$ (%) | P value |
| Design (cohort vs. RCT) | 0.00 | 0.916 | 0.00 | 0.78 | 82.84 | 0.62 | 0.00 | 0.716 |
| Assay (ELISA vs. other) | 0.00 | 0.753 | 84.97 | 0.07 | 0.00 | 0.05 | 0.00 | 0.435 |
| Sample (Serum vs. plasma) | 0.00 | 0.56 | 0.00 | 0.557 | 19.2 | 0.662 | 0.00 | 0.716 |
| IPF consensus (2011 vs. other) | N/A | N/A | N/A | N/A | N/A | N/A | N/A | N/A |
| Centre (single vs. multi) | N/A | N/A | N/A | N/A | N/A | N/A | N/A | N/A |
| Publication type (peer reviewed) | N/A | N/A | N/A | N/A | N/A | N/A | N/A | N/A |

Supplementary Table 2 **-** Results of meta-regression for variables assessed separated by study outcomes. Sample sizes for each outcome shown (n). $R^2$ and p values from meta-regression shown where applicable.
N/A, not applicable.

| Author (year) | Sample size | Follow up (months) | Effect size (Variance) | Level of adjustment | Effect size reported for |
|---|---|---|---|---|---|
| **MMP-7 (IPD unavailable)** | | | | | |
| Sokai (2015) | 57 | 15 | Not significant (NR) | NR | NR |
| Peljto (2013) | 438 | 19 | 2.18 (95% CI 1.1-4.32) | b,d,e,h | bio > or < 5.7ng/mL |
| **SP-A** | | | | | |
| Kinder (2009) | 82 | 36 | HR 3.27 (95% CI 1.49-7.17) | a,b,c,d,e,g | per bio SD |
| Doubkova (2016) | 18 | NR | Not significant (NR) | x | bio > or < median (98.1ng/mL) |
| Hamai (2016) | 65 | 31 | HR 1.01 (95% CI 0.99-1.02) | x | continuous |
| **SP-D** | | | | | |
| Kinder (2009) | 82 | 36 | HR 2.04 (95% CI 0.99-4.22) | a,b,c,d,e,g | per bio SD |
| Collard (2010) | 67 | NR | OR 1.23 (95% CI 0.36-4.21) | "Bivariate" - NR | log change in bio |
| Doubkova (2016) | 18 | NR | Not significant (NR) | x | bio > or < median (623.1ng/mL) |
| Hamai (2016) | 65 | 31 | HR 1.00 (95% CI 0.99-1.002) | x | continuous |
| Maher (2017) - *Validation* | 206 | 36 | HR 2.72 (95% CI 1.65-4.48) | x | bio > or < 38.7ng/mL |
| **CCL-18** | | | | | |
| Prasse (2009) | 72 | 24 | HR 7.98 (95% CI 2.49-25.51) | a,b,c,d,e | bio > or < 150ng/mL |
| Hamai (2016) | 65 | 31 | HR 1.007 (95% CI 0.99-1.01) | X | continuous |
| Neighbors (2018) – *Test* | 123 | 12 | OR 4.4 (95% CI 1.13-17.15) | x | bio ≥ or < median |
| Neighbors (2018) – *Replication* | 237 | 12 | OR 3.37 (95% CI 1.17-9.67) | x | bio ≥ or < median |
| **CXCL-13** | | | | | |
| Guo (2020) | 126 | 60 | HR 1.03 (95% CI 1.02-1.06) | a | bio > or < 62pg/mL |

| | | | | | |
|---|---|---|---|---|---|
| Vuga (2014) | 95 | >24 | HR 14.9 (95% CI 1.1-197.2) | a,b,d,e | bio > or < highest quartile |
| Neighbors (2018) – *Test* | 123 | 12 | OR 2.95 (95% CI 0.76-11.46) | x | bio ≥ or < median |
| Neighbors (2018) – *Replication* | 237 | 12 | OR 6.17 (95% CI 1.75-21.8) | x | bio ≥ or < median |
| **KL-6** | | | | | |
| Collard (2010) | 67 | NR | OR 0.41 (95% CI 0.06-2.93) | "Bivariate" - NR | bio log change |
| Hamai (2016) | 65 | 31 | HR 1.001 (95% CI 1.00-1.002) | a,b,c | continuous |
| Guo (2020) | 126 | 60 | HR 1.83 (95% CI 1.01-3.69) | a | bio > or < 800U/mL |
| **IL-8** | | | | | |
| Richards (2012) – *Derivation* | 140 | 22 | HR 2.4 (95% CI 1.2-4.79) | a,b,d | bio > or < 0.0029 |
| Richards (2012) – *Validation* | 101 | 17 | HR 2.3 (95% CI 0.94-5.64) | a,b,d | bio > or < 0.0097 |
| Papiris (2018) | 41 | 12 | OR 1.067 (95% CI 1.01-1.12) | x | per increase of 1pg/mL |
| **CA19-9** | | | | | |
| Maher (2017) – *Validation* | 206 | 36 | HR 2.95 (95% CI 1.82-4.78) | x | bio > or < 22 U/mL |
| **CA-125** | | | | | |
| Maher (2017) – *Validation* | 206 | 36 | HR 3.01 (95% CI 1.64-5.54) | x | bio > or < 12 U/mL |
| **LOXL2** | | | | | |
| Chien (2014) – *ARTEMIS* | 69 | 24 | HR 1.87 (95% CI 0.28-12.45) | d,e,f,h | bio > or ≤ 800pg/mL |
| Chien (2014) – *GAP* | 104 | 24 | HR 2.28 (95% CI 1.18-4.38) | b | bio > or ≤ 700pg/mL |
| **Periostin** | | | | | |
| Okamoto (2011) | 77 | 36 | Not significant (NR) | x | NR |
| Neighbors (2018) - *Test* | 123 | 12 | OR 3.05 (95% CI 0.79-11.88) | x | bio ≥ or < median |
| Neighbors (2018) – *Replication* | 237 | 12 | OR 1.91 (95% CI 0.72-5.05) | x | bio ≥ or < median |
| **YKL-40** | | | | | |

| | | | | | |
|---|---|---|---|---|---|
| Neighbors (2018) – *Test* | 123 | 12 | OR 1.77 (95% CI 0.53-5.92) | x | bio ≥ or < median |
| Neighbors (2018) – *Replication* | 237 | 12 | OR 2.7 (95% CI 0.94-7.75) | x | bio ≥ or < median |
| **ICAM-1** | | | | | |
| Richards (2012) - *Derivation* | 140 | 22 | HR 2.6 (95% CI 1.43-4.73) | a,b,d | bio > or < 202.5ng/mL |
| Richards (2012) – *Validation* | 101 | 17 | HR 2.8 (95% CI 1.36-5.76) | a,b,d | bio > or < 300ng/mL |
| **ECM neoepitopes** | | | | | |
| Jenkins (2015) –*Discovery* ***BGM*** | 55 | 26 | HR 1.17 (95% CI 0.53-2.58) | x | two-fold increase in bio value |
| Jenkins (2015) –*Validation* ***BGM*** | 134 | 21 | HR 1.34 (95% CI 0.92-1.97) | x | two-fold increase in bio value |
| Jenkins (2015) –*Discovery* ***C1M*** | 55 | 26 | HR 1.21 (95% CI 0.66-2.22) | x | two-fold increase in bio value |
| Jenkins (2015) –*Validation* ***C1M*** | 134 | 21 | HR 1.62 (95% CI 1.14-2.31) | x | two-fold increase in bio value |
| Jenkins (2015) –*Discovery* ***C3A*** | 55 | 26 | HR 1.34 (95% CI 0.95-1.88) | x | two-fold increase in bio value |
| Jenkins (2015) –*Validation* ***C3A*** | 134 | 21 | HR 1.91 (95% CI 1.06-3.46) | x | two-fold increase in bio value |
| Jenkins (2015) –*Discovery* ***C3M*** | 55 | 26 | HR 2.18 (95% CI 0.95-5.00) | x | two-fold increase in bio value |
| Jenkins (2015) –*Validation* ***C3M*** | 134 | 21 | HR 1.56 (95% CI 0.94-2.59) | x | two-fold increase in bio value |
| Jenkins (2015) –*Discovery* ***C5M*** | 55 | 26 | HR 1.66 (95% CI 0.95-2.91) | x | two-fold increase in bio value |
| Jenkins (2015) –*Validation* ***C5M*** | 134 | 21 | HR 1.07 (95% CI 0.66-1.72) | x | two-fold increase in bio value |
| Jenkins (2015) –*Discovery* ***C6M*** | 55 | 26 | HR 1.49 (95% CI 0.86-2.56) | x | two-fold increase in bio value |
| Jenkins (2015) –*Validation* ***C6M*** | 134 | 21 | HR 1.39 (95% CI 0.93-2.06) | x | two-fold increase in bio value |
| Jenkins (2015) –*Discovery* ***CRPM*** | 55 | 26 | HR 3.74 (95% CI 1.46-9.58) | x | two-fold increase in bio value |
| Jenkins (2015) –*Validation* ***CRPM*** | 134 | 21 | HR 1.87 (95% CI 0.98-3.56) | x | two-fold increase in bio value |
| Jenkins (2015) – *Discovery* ***ELM*** | 55 | 26 | HR 0.96 (95% CI 0.48-1.92) | x | two-fold increase in bio value |
| Jenkins (2015) – *Discovery* ***ELM2*** | 55 | 26 | HR 0.96 (95% CI 0.75-1.24) | x | two-fold increase in bio value |

| | | | | | |
|---|---|---|---|---|---|
| Jenkins (2015) – *Discovery **P3NP*** | 55 | 26 | HR 1.48 (95% CI 0.67-3.27) | x | two-fold increase in bio value |
| Jenkins (2015) – *Discovery **VICM*** | 55 | 26 | HR 1.11 (95% CI 0.83-1.49) | x | two-fold increase in bio value |
| **Collagen synthesis peptides** | | | | | |
| Organ (2019) **P1NP** | 145 | 34 | HR 0.81 (95% CI 0.6-1.11) | d,e | two-fold increase in bio value |
| Organ (2019) **PRO-C3** | 145 | 34 | HR 1.2 (95% CI 0.74-1.93) | d,e | two-fold increase in bio value |
| Hoyer (2020) **PRO-C3** | 184 | 36 | HR 2.32 (95% CI 1.33-4.04) | a | continuous |
| Organ (2019) **PRO-C6** | 145 | 34 | HR 1.11 (95% CI 0.57-2.16) | d,e | two-fold increase in bio value |
| Hoyer (2020) **PRO-C6** | 184 | 36 | HR 2.18 (95% CI 0.74-4.35) | a | continuous |
| Organ (2019) **P1NP:C1M** | 145 | 34 | HR 0.77 (95% CI 0.6-0.99) | d,e | two-fold increase in bio value |
| Organ (2019) **PRO-C3:C3M** | 145 | 34 | HR 1.17 (95% CI 0.77-1.79) | d,e | two-fold increase in bio value |
| Organ (2019) **PRO-C6:C6M** | 145 | 34 | HR 0.86 (95% CI 0.59-1.26) | d,e | two-fold increase in bio value |
| Hoyer (2020) **PRO-C6** | 184 | 36 | HR 1.8 (95% CI 0.74-4.35) | a | continuous |

Supplementary Table 3 – Studies reporting mortality outcomes
x=no adjustments, a=age, b=gender, c=smoking, d=FVC e=DLCO, f= 6MWT, g=race, h=medication

bio, biomarker; HR, hazard ratio; IPD, individual participant data; NR, not reported; OR, odds ratio

| Author (year) | Sample size | Follow up (months) | Effect size (Variance) | Level of adjustment | Effect size reported for |
|---|---|---|---|---|---|
| **SP-D** | | | | | |
| Maher (2017) - *Discovery* | 106 | 36 | HR 1.01 (95% CI 0.97-1.06) | x | rising vs stable bio over 3 months |
| Maher (2017) – *Validation* | 206 | 36 | HR 0.99 (95% CI 0.59-1.67) | a,b,c,d | rising vs stable bio over 3 months |
| **CA19-9** | | | | | |
| Maher (2017) - *Discovery* | 106 | 36 | HR 1.02 (95% CI 1.00-1.05) | X | rising vs stable bio over 3 months |
| Maher (2017) – *Validation* | 206 | 36 | HR 1.39 (95% CI 0.79-2.46) | a,b,c,d | rising vs stable bio over 3 months |
| **CA-125** | | | | | |
| Maher (2017) - *Discovery* | 106 | 36 | HR 1.77 (95% CI 1.39-2.26) | x | rising vs stable bio over 3 months |
| Maher (2017) – *Validation* | 206 | 36 | HR 2.39 (95% CI 1.4-4.08) | a,b,c,d | rising vs stable bio over 3 months |
| **ICAM-1** | | | | | |
| Maher (2017) - *Discovery* | 106 | 36 | HR 1.002 (95% CI 0.99-1.01) | x | rising vs stable bio over 3 months |
| **IGFBP-2** | | | | | |
| Maher (2017) - *Discovery* | 106 | 36 | HR 1.02 (95% CI 1.002-1.03) | x | rising vs stable bio over 3 months |
| **IL-8** | | | | | |
| Maher (2017) - *Discovery* | 106 | 36 | HR 1.02 (95% CI 0.98-1.07) | x | rising vs stable bio over 3 months |
| **ECM neoepitopes** | | | | | |
| Jenkins (2015) –*Validation **BGM*** | 134 | 21 | HR 1.07 (95% CI 1.00-1.15) | a,c,d,e | rising vs stable bio over 3 months |
| Organ (2019) **BGM** | 145 | 34 | HR 1.41 (95% CI 0.8-2.47) | a,b,c | rising vs stable bio over 3 months |
| Jenkins (2015) –*Validation **C1M*** | 134 | 21 | HR 1.01 (95% CI 1.00-1.02) | a,c,d,e | rising vs stable bio over 3 months |
| Organ (2019) **C1M** | 145 | 34 | HR 1.84 (95% CI 1.03-3.27) | a,b,c | rising vs stable bio over 3 months |

| | | | | | |
|---|---|---|---|---|---|
| Jenkins (2015) –*Validation* ***C3A*** | 134 | 21 | HR 1.05 (95% CI 1.01-1.1) | a,c,d,e | rising vs stable bio over 3 months |
| Jenkins (2015) –*Validation* ***C3M*** | 134 | 21 | HR 1.1 (95% CI 1.04-1.17) | a,c,d,e | rising vs stable bio over 3 months |
| Organ (2019) **C3M** | 145 | 34 | HR 2.44 (95% CI 1.39-4.31) | a,b,c | rising vs stable bio over 3 months |
| Jenkins (2015) –*Validation* ***C5M*** | 134 | 21 | HR 1.00 (95% CI 1.00-1.00) | a,c,d,e | rising vs stable bio over 3 months |
| Jenkins (2015) –*Validation* ***C6M*** | 134 | 21 | HR 1.04 (95% CI 1.01-1.08) | a,c,d,e | rising vs stable bio over 3 months |
| Organ (2019) **C6M** | 145 | 34 | HR 2.19 (95% CI 1.25-3.82) | a,b,c | rising vs stable bio over 3 months |
| Jenkins (2015) –*Validation* ***CRPM*** | 134 | 21 | HR 1.33 (95% CI 1.1-1.6) | a,c,d,e | rising vs stable bio over 3 months |
| Organ (2019) **CRPM** | 145 | 34 | HR 2.13 (95% CI 1.21-3.75) | a,b,c | rising vs stable bio over 3 months |
| Jenkins 2015) – *Validation* ***VICM*** | 55 | 26 | HR 1.01 (95% CI 0.99-1.03) | a,c,d,e | rising vs stable bio over 3 months |
| **Collagen synthesis peptides** | | | | | |
| Organ (2019) **P1NP** | 145 | 34 | HR 0.76 (95% CI 0.44-1.3) | a,b,c | rising vs stable bio over 3 months |
| Organ (2019) **PRO-C3** | 145 | 34 | HR 1.62 (95% CI 0.95-2.79) | a,b,c | rising vs stable bio over 3 months |
| Organ (2019) **PRO-C6** | 145 | 34 | HR 1.14 (95% CI 0.67-1.93) | a,b,c | rising vs stable bio over 3 months |
| Organ (2019) **P1NP:C1M** | 145 | 34 | HR 0.73 (95% CI 0.41-1.29) | a,b,c | rising ratio levels |
| Organ (2019) **PRO-C3:C3M** | 145 | 34 | HR 0.83 (95% CI 0.49-1.43) | a,b,c | rising ratio levels |
| Organ (2019) **PRO-C6:C6M** | 145 | 34 | HR 0.55 (95% CI 0.32-0.95) | a,b,c | rising ratio levels |

Supplementary Table 4 – Studies reporting short term biomarkers change and their association with mortality

x=no adjustments, a=age, b=gender, c=smoking, d=FVC e=DLCO, f= 6MWT, g=race, h=medication
bio, biomarker; HR, hazard ratio.

| Author (year) | Sample size | Timepoint of outcome (months) | Disease progression definition | Effect size (Variance) | Level of adjustment | Effect size reported for |
|---|---|---|---|---|---|---|
| **MMP-7 (IPD unavailable)** | | | | | | |
| Sokai (2015) | 57 | 6 | FVC decline ≥10% or $DL_{CO}$ ≥ 15% decline or respiratory failure or death | Not significant (NR) | NR | NR |
| Bauer (2017) | 211 | 19 | FVC decline ≥ 10% or $DL_{CO}$ ≥ 15% decline or respiratory failure or death | HR 2.2 (95% CI 1.4-3.7) | NR | bio < or ≥ 3.8ng/mL |
| **SP-A** | | | | | | |
| Raghu (2018) | 130 | 12 | FVC decrease ≥10% predicted or $DL_{CO}$ decrease > 15% or lung transplantation or death | AUROC 0.61 (90% CI 0.52-0.7) | NR | NR |
| **SP-D** | | | | | | |
| Collard (2010) | 67 | NR | Acute exacerbation | 361ng/mL vs 294ng/mL (p=0.01) | x | median bio in event and non-event group |
| Maher (2017) *Discovery* | 104 | 12 | All-cause mortality or FVC decline ≥ 10% | GR 1.35 (95% CI 1.1-1.649) | x | bio level in progressive vs. stable group |
| Maher (2017) *Validation* | 204 | 12 | All-cause mortality or FVC decline ≥ 10% | GR 1.35 (95% CI 1.12-1.62) | x | bio level in progressive vs. stable group |
| Raghu (2018) | 130 | 12 | FVC decrease ≥10% predicted or $DL_{CO}$ decrease > 15% or lung transplantation or death | AUROC 0.62 (90% CI 0.53-0.7) | NR | NR |
| **CCL-18** | | | | | | |
| Prasse (2009) | 67 | 24 | FVC decline ≥ 10% prediced or death | OR 6.75 (95% CI 2.52-18.1) | x | bio < or > 150ng/mL |
| Ohshimo (2014) | 77 | 36 | Acute exacerbation | HR 2.92 (95% CI 0.76-11.4) | x | bio > or < 212ng/mL |
| Neighbors (2018) *Test* | 123 | 12 | FVC ≥10% absolute decline, 50m decline in 6MWT or death | HR 1.64 (95% CI 1.04-2.83) | x | 'high' vs 'low' bio |
| Neighbors (2018) *Replication* | 237 | 12 | FVC ≥10% absolute decline, 50m decline in 6MWT or death | HR 1.32 (95% CI 0.76-2.13) | x | 'high' vs 'low' bio |
| Raghu (2018) | 130 | 12 | FVC decrease ≥10% predicted or $DL_{CO}$ decrease > 15% or lung transplantation or death | AUROC 0.62 (90% CI 0.54-0.71) | NR | bio > or < 150ng/mL |

| | | | | | | |
|---|---|---|---|---|---|---|
| **CXCL-13** | | | | | | |
| Neighbors (2018) *Test* | 123 | 12 | FVC ≥10% absolute decline, 50m decline in 6MWT or death | HR 1.23 (95% CI 0.89-1.69) | x | ‘high’ vs ‘low’ bio |
| Neighbors (2018) *Replication* | 237 | 12 | FVC ≥10% absolute decline, 50m decline in 6MWT or death | Not significant (NR) | x | ‘high’ vs ‘low’ bio |
| **KL-6** | | | | | | |
| Collard (2010) | 67 | NR | Acute exacerbation | 1791 U/mL vs 895 U/mL (p=0.003) | x | median bio in event and non-event group |
| Ohshimo (2014) | 77 | 36 | Acute exacerbation | HR 11.8 (95% CI 1.43-97.8) | a,b,c,h | bio > or < 1300U/mL |
| Jiang (2018) | 20 | 12 | FVC decline ≥ 10% or $DL_{CO}$ decline ≥ 15%, or death | OR 1.00 (95% CI 1.00-1.00) | x | continuous bio |
| Raghu (2018) | 130 | 12 | FVC decrease ≥10% predicted or $DL_{CO}$ decrease > 15% or lung transplantation or death | AUROC 0.6 (90% CI 0.51-0.68) | NR | NR |
| **IL-8** | | | | | | |
| Richards (2012) *Derivation* | 140 | 12 | FVC relative decline ≥ 10% | HR 2.00 (95% CI 1.22-3.28) | a,b,d | bio > or < 0.0092ng/mL |
| Richards (2012) *Validation* | 101 | 12 | FVC relative decline ≥ 10% | HR 1.2 (95% CI 0.5-2.85) | a,b,d | bio > or < 0.0092ng/mL |
| Maher (2017) *Discovery* | 104 | 12 | All-cause mortality or FVC decline ≥ 10% | GR 1.51 (95% CI 1.12-2.023) | x | bio level in progressive vs. stable group |
| **CA19-9** | | | | | | |
| Maher (2017) *Discovery* | 104 | 12 | All-cause mortality or FVC decline ≥ 10% | GR 3.12 (95% CI 1.7-5.7) | x | bio level in progressive vs. stable group |
| Maher (2017) *Validation* | 204 | 12 | All-cause mortality or FVC decline ≥ 10% | GR 2.42 (95% CI 1.6-3.65) | x | bio level in progressive vs. stable group |
| **CA125** | | | | | | |
| Maher (2017) *Discovery* | 104 | 12 | All-cause mortality or FVC decline ≥ 10% | Not significant (NR) | x | bio level in progressive vs. stable group |
| Maher (2017) *Validation* | 204 | 12 | All-cause mortality or FVC decline ≥ 10% | GR 1.26 (95% CI 1.05-1.51) | x | bio level in progressive vs. stable group |

| | | | | | | |
|---|---|---|---|---|---|---|
| **LOXL2** | | | | | | |
| Chien (2014) *ARTEMIS* | 69 | 24 | Mortality, hospitalisation or lung function decline (FVC≥10% & $DL_{CO}$≥5%, or $DL_{CO}$ ≥ 15% and FVC≥5%) | HR 5.41 (95% CI 1.65-17.73) | d,e,f,h | bio > or ≤ 800pg/mL |
| Chien (2014) *GAP* | 70 | 24 | Mortality, hospitalisation or lung function decline (FVC≥10% & $DL_{CO}$≥5%, or $DL_{CO}$ ≥ 15% and FVC≥5%) | HR 1.78 (95% CI 1.01-3.11) | x | bio > or ≤ 700pg/mL |
| **Periostin** | | | | | | |
| Naik (2012) | 50 | 11 | Death, acute exacerbation, transplantation, relative FVC decline ≥ 10% or $DL_{CO}$ > 15% | HR 1.47 (95% CI 1.03-2.1) | a,b,c,d,e | per bio SD |
| Neighbors (2018) *Test* | 123 | 12 | FVC ≥10% absolute decline, 50m decline in 6MWT or death | HR 2.08 (95% CI 1.24-3.47) | x | 'high' vs 'low' bio |
| Neighbors (2018) *Replication* | 237 | 12 | FVC ≥10% absolute decline, 50m decline in 6MWT or death | HR 1.75 (95% CI 0.87-2.84) | x | 'high' vs 'low' bio |
| Raghu (2018) | 130 | 12 | FVC decrease ≥10% predicted or $DL_{CO}$ decrease > 15% or lung transplantation or death | AUROC 0.6 (90% CI 0.51-0.69) | NR | NR |
| **YKL-40** | | | | | | |
| Neighbors (2018) *Test* | 123 | 12 | FVC ≥10% absolute decline, 50m decline in 6MWT or death | HR 1.39 (95% CI 0.79-2.41) | x | 'high' vs 'low' bio |
| Neighbors (2018) *Replication* | 237 | 12 | FVC ≥10% absolute decline, 50m decline in 6MWT or death | Not significant (NR) | x | 'high' vs 'low' bio |
| Raghu (2018) | 130 | 12 | FVC decrease ≥10% predicted or $DL_{CO}$ decrease > 15% or lung transplantation or death | AUROC 0.58 (90% CI 0.49-0.67) | NR | NR |
| ICAM-1 | | | | | | |
| Richards (2012) *Derivation* | 140 | 12 | FVC relative decline ≥ 10% | HR 1.6 (95% CI 1.00-2.56) | a,b,d | bio > or < 202.5ng/mL |
| Richards (2012) *Validation* | 101 | 12 | FVC relative decline ≥ 10% | HR 2.2 (95% CI 1.21-4.01) | a,b,d | bio > or < 262ng/mL |
| Maher (2017) *Discovery* | 104 | 12 | All-cause mortality or FVC decline ≥ 10% | GR 1.29 (95% CI 1.02-1.65) | x | bio level in progressive vs. stable group |
| Raghu 2018 | 130 | 12 | FVC decrease ≥10% predicted or $DL_{CO}$ decrease > 15% or lung transplantation or death | AUROC 0.65 (90% CI 0.56-0.73) | NR | NR |
| **ECM neoepitopes** | | | | | | |

| | | | | | | |
|---|---|---|---|---|---|---|
| Jenkins (2015)<br>*D+V cohort* **BGM** | 186 | 12 | All-cause mortality or FVC decline ≥ 10% | Not significant (NR) | x | bio level in progressive vs. stable group |
| Jenkins (2015)<br>*D+V cohort* **C1M** | 186 | 12 | All-cause mortality or FVC decline ≥ 10% | Not significant (NR) | x | bio level in progressive vs. stable group |
| Jenkins (2015)<br>*D+V cohort* **C3M** | 186 | 12 | All-cause mortality or FVC decline ≥ 10% | P=0.011 (NR) | x | bio level in progressive vs. stable group |
| Jenkins (2015)<br>*D+V cohort* **C5M** | 186 | 12 | All-cause mortality or FVC decline ≥ 10% | Not significant (NR) | x | bio level in progressive vs. stable group |
| Jenkins (2015)<br>*D+V cohort* **C6M** | 186 | 12 | All-cause mortality or FVC decline ≥ 10% | P=0.013 (NR | x | bio level in progressive vs. stable group |
| Jenkins (2015)<br>*D+V cohort* **CRPM** | 186 | 12 | All-cause mortality or FVC decline ≥ 10% | P=0.014 (NR) | x | bio level in progressive vs. stable group |
| Jenkins (2015)<br>*D+V cohort* **VICM** | 186 | 12 | All-cause mortality or FVC decline ≥ 10% | P=0.033 (NR) | x | bio level in progressive vs. stable group |
| Jenkins (2015)<br>*D+V cohort* **C3A** | 186 | 12 | All-cause mortality or FVC decline ≥ 10% | P=0.003 (NR) | x | bio level in progressive vs. stable group |
| Jenkins (2015)<br>*Discovery only* **P3NP** | 186 | 12 | All-cause mortality or FVC decline ≥ 10% | P=0.63 (NR) | x | bio level in progressive vs. stable group |
| Jenkins (2015)<br>*Discovery only* **ELM** | 186 | 12 | All-cause mortality or FVC decline ≥ 10% | P=0.55 (NR) | x | bio level in progressive vs. stable group |
| Jenkins (2015)<br>*Discovery only* **ELM2** | 186 | 12 | All-cause mortality or FVC decline ≥ 10% | P=0.42 (NR) | x | bio level in progressive vs. stable group |
| Hoyer (2020)<br>**PROC3** | 184 | 6 | All-cause mortality or FVC decline ≥ 10% | P=0.005 (NR) | NR | NR |
| Hoyer (2020)<br>**PROC6** | 184 | 6 | All-cause mortality or FVC decline ≥ 10% | P=0.031 (NR) | NR | NR |

Supplementary Table 5 – Studies reporting disease progression outcomes including definition of disease progression outcome used and effect sizes reported.

x=no adjustments, a=age, b=gender, c=smoking, d=FVC e=$DL_{CO}$, f= 6MWT, g=race, h=medication, NR=not reported

bio, biomarker; AUROC; area under the receiver operating characteristics; $DL_{CO,}$ gas transfer for carbon monoxide; FVC, forced vital capacity; GR, group ratio; HR, hazard ratio; IPD, individual participant data; NR, not reported; OR, odds ratio; 6MWT, 6-minute walk test;

| Author (year) | Sample size | Timepoint of outcome (months) | Disease progression definition | Effect size (Variance) | Level of adjustment | Effect size reported for |
|---|---|---|---|---|---|---|
| **MMP-7 (IPD unavailable)** | | | | | | |
| Bauer et al (2017) | 211 | "Study period" | FVC ≥10% decline, $DL_{CO}$ ≥ 15%, acute exacerbation or death | OR 1.9 (95% CI 1.2-3.0) | NR | Two-fold change in bio over 4 months |
| **SP-D** | | | | | | |
| Maher et al (2017) *Discovery* | 106 | 12 | All-cause mortality or FVC decline ≥ 10% | p=0.029 | x | rising vs stable bio over 3 months |
| Maher et al (2017) *Validation* | 206 | 12 | All-cause mortality or FVC decline ≥ 10% | Not significant (NR) | x | rising vs stable bio over 3 months |
| **CXCL-13** | | | | | | |
| Vuga et al (2014) | 95 | >24 | Respiratory failure | HR 7.2 (95% CI 1.3-40.0) | x | bio "increase greatest vs. less increased" (time not specified) |
| **CA19-9** | | | | | | |
| Maher et al (2017) *Discovery* | 106 | 12 | All-cause mortality or FVC decline ≥ 10% | p<0.001 | x | rising vs stable bio over 3 months |
| Maher et al (2017) *Validation* | 206 | 12 | All-cause mortality or FVC decline ≥ 10% | Not significant (NR) | x | rising vs stable bio over 3 months |
| **CA125** | | | | | | |
| Maher et al (2017) *Discovery* | 106 | 12 | All-cause mortality or FVC decline ≥ 10% | p=0.041 | x | rising vs stable bio over 3 months |
| Maher et al (2017) *Validation* | 206 | 12 | All-cause mortality or FVC decline ≥ 10% | p=0.0028 | x | rising vs stable bio over 3 months |
| **KL-6** | | | | | | |
| Jiang et al (2018) | 20 | 12 | FVC decline ≥ 10%, $DL_{CO}$ decline ≥ 15% or death | OR 3.61 (95% CI 1.05-6.22) | a,b,c,d,e | Change in KL-6 (not otherwise specified) |

Supplementary Table 6 – Studies reporting short term biomarkers change and their association with disease progression

x=no adjustments, a=age, b=gender, c=smoking, d=FVC e=$DL_{CO}$, f= 6MWT, g=race, h=medication, NR=not reported
bio, biomarker; $DL_{CO,}$ gas transfer for carbon monoxide; FVC, forced vital capacity; GR, group ratio; HR, hazard ratio; IPD, individual participant data; NR, not reported; OR, odds ratio

| Author (year) | Sample size | FVC change measured at (months) | Effect size (Variance) | Level of adjustment | Effect size reported for |
|---|---|---|---|---|---|
| **MMP-7 (IPD unavailable)** | | | | | |
| Bauer (2017) | 195 | 4 | p=0.004 (NR) | x | baseline bio correlation with %pred FVC change |
| **SP-A** | | | | | |
| Doubkova (2016) | 18 | NR | 155.8 ng/mL vs 87.15 ng/mL; p=0.01 | x | baseline bio in PFT “improvement” vs “stabilisation” |
| **SP-D** | | | | | |
| Doubkova (2016) | 18 | NR | 861.4ng/mL vs. 802.8ng/mL; p=0.76 | x | baseline bio in PFT “improvement” vs “stabilisation” |
| Kennedy (2015) | 13 | 6 | r= -0.64 (95% CI -0.89 to -0.08) | x | baseline bio correlation with %pred FVC change |
| Ohta (2017) | 60 | 6-12 | r= 0.09 (p>0.05) | x | baseline bio correlation with %pred FVC change |
| **CCL-18** | | | | | |
| Neighbors (2018) – *Test* | 123 | 12 | -3.1% (p=0.03) | x | %pred FVC change in baseline bio ≥ or < median (411.5ng/mL) |
| Neighbors (2018) – *Replication* | 237 | 12 | -3.6% (p=0.004) | x | %pred FVC change in baseline bio ≥ or < median (458.6ng/mL) |
| Prasse (2009) | 67 | 6 | r=0.54 (p<0.0001) | x | baseline bio correlation with %pred FVC change |
| **CXCL-13** | | | | | |
| Guo (2020) | 126 | 12 | r= 0.56 (p<0.001) | x | baseline bio correlation with %pred FVC change |
| Neighbors (2018) – *Test* | 123 | 12 | -3.2% (p=0.06) | x | %pred FVC change in baseline bio ≥ or < median (87.9ng/mL) |
| Neighbors (2018) – *Replication* | 237 | 12 | -3.7% (p=0.05) | x | %pred FVC change in baseline bio ≥ or < median (88.7ng/mL) |
| **KL-6** | | | | | |
| Guo (2020) | 126 | 12 | r= 0.71 (p<0.001) | x | baseline bio correlation with %pred FVC change |
| Ohta (2017) | 60 | 6-12 | r= 0.09 (p>0.05) | x | baseline bio correlation with %pred FVC change |

| | | | | | |
|---|---|---|---|---|---|
| Okamoto (2011) | 26 | 6 | Not significant (NR) | x | baseline bio correlation with %pred FVC change |
| **Periostin** | | | | | |
| Neighbors (2018) – *Test* | 123 | 12 | -3.6% (p<0.001) | x | %pred FVC change in baseline bio ≥ or < median (67.8ng/mL) |
| Neighbors (2018) – *Replication* | 237 | 12 | -2.5% (p=0.19) | x | %pred FVC change in baseline bio ≥ or < median (65.4ng/mL) |
| Ohta (2017) | 60 | 6-12 | r= -0.43 (p<0.01) | x | baseline bio correlation with %pred FVC change |
| Okamoto (2011) | 26 | 6 | r= -0.50 (p<0.01) | x | baseline bio correlation with %pred FVC change |
| **YKL-40** | | | | | |
| Neighbors (2018) – *Test* | 123 | 12 | -2.4% (p=0.04) | x | %pred FVC change in baseline bio ≥ or < median (100.3ng/mL) |
| Neighbors (2018) – *Replication* | 237 | 12 | -1.5% (p=0.70) | x | %pred FVC change in baseline bio ≥ or < median (109.5ng/mL) |

Supplementary Table 7 – Studies reporting association with baseline biomarkers and change in forced vital capacity (FVC).
bio, biomarker; x = no adjustments

IPD, individual participant data.

| Outcome | The GRADE domains | Ratings for quality of evidence |
|---|---|---|
| **Baseline MMP-7** | | |
| Overall mortality (10 studies; 1492 participants) | Risk of bias | All studies included well-defined participants diagnosed according to international consensus guidelines. Exposure and outcomes were measured objectively and consistently for all participants. Covariates were adjusted for using IPD. Follow up was sufficiently long to enable outcomes to occur. |
| | Imprecision | Effect sizes in most studies favour MMP-7 as a marker of mortality. |
| | Inconsistency | Substantial heterogeneity not explained by variability in the factors assessed |
| | Indirectness | No serious indirectness. All patients had IPF according to consensus criteria and were untreated. MMP-7 was measured at baseline in all studies, and overall mortality measured from IPD. |
| | Publication bias | No publication bias as indicated by funnel plots and Egger's tests |
| | Certainty of evidence | Moderate certainty of evidence |
| 12-month mortality (10 studies; 1492 participants) | Risk of bias | All studies included well-defined participants diagnosed according to international consensus guidelines. Exposure and outcomes were measured objectively and consistently for all participants. Covariates were adjusted for using IPD. Follow up was sufficiently long to enable outcomes to occur. |
| | Imprecision | Imprecision present with wide confidence interval of 0.99-1.78. |
| | Inconsistency | Substantial heterogeneity not explained by variability in the factors assessed |
| | Indirectness | No serious indirectness. All patients had IPF according to consensus criteria and were untreated. MMP-7 was measured at baseline in all studies, and 12-month mortality measured from IPD. |
| | Publication bias | No publication bias as indicated by funnel plots and Egger's tests |

| | | |
|---|---|---|
| | Certainty of evidence | Moderate certainty of evidence |
| Disease progression (10 studies; 1383 participants) | Risk of bias | All studies included well-defined participants diagnosed according to international consensus guidelines. Exposure was measured objectively and consistently for all participants. Disease progression definition was standardised. Covariates were adjusted for using IPD. Follow up was sufficiently long to enable outcomes to occur. |
| | Imprecision | Effect sizes consistently favour MMP-7 as a prognostic marker, although confidence intervals commonly cross 1. Overall estimate has appropriately narrow confidence interval supporting MMP-7 as a biomarker of disease progression. |
| | Inconsistency | No heterogeneity demonstrated. |
| | Indirectness | No serious indirectness. All patients had IPF according to consensus criteria and were untreated. MMP-7 was measured at baseline in all studies, and disease progression standardised using IPD. |
| | Publication bias | No publication bias as indicated by funnel plots and Egger's tests |
| | Certainty of evidence | High certainty of evidence. |
| Change in FVC at 12 months (8 studies; 891 participants) | Risk of bias | All studies included well-defined participants diagnosed according to international consensus guidelines. Exposure was measured objectively and consistently for all participants. Change in FVC was measured objectively and consistently for all participants. Covariates were adjusted for using IPD. Follow up was sufficiently long to enable outcomes to occur. |
| | Imprecision | The majority of the studies show MMP-7 to result in a negative change in FVC at 12 months, although confidence intervals cross 0 in all individual studies. Overall confidence interval does not cross 0. |
| | Inconsistency | No evidence of heterogeneity |
| | Indirectness | No serious indirectness. All patients had IPF according to consensus criteria and were untreated. MMP-7 was measured at baseline in all studies and change in FVC standardised using IPD. |
| | Publication bias | No obvious funnel plot asymmetry, although unable to assess statistically due to small number of studies |
| | Certainty of evidence | High certainty of evidence. |

| Three-month MMP-7 change | | |
|---|---|---|
| Overall mortality (4 studies; 498 participants) | Risk of bias | All studies included well-defined participants diagnosed according to international consensus guidelines. Exposure and outcomes were measured objectively and consistently for all participants. Covariates were adjusted for using IPD. Follow up was sufficiently long to enable outcomes to occur. |
| | Imprecision | Wide confidence intervals in individual studies but narrow confidence interval for overall effect size (no effect) |
| | Inconsistency | Substantial heterogeneity not explained by variability in the factors assessed |
| | Indirectness | No serious indirectness. All patients had IPF according to consensus criteria and were untreated. MMP-7 was measured at baseline in all studies, and overall mortality measured from IPD. |
| | Publication bias | No obvious funnel plot asymmetry, although unable to assess statistically due to small number of studies |
| | Certainty of evidence | Moderate certainty of evidence |
| 12-month mortality (4 studies; 498 participants) | Risk of bias | All studies included well-defined participants diagnosed according to international consensus guidelines. Exposure and outcomes were measured objectively and consistently for all participants. Covariates were adjusted for using IPD. Follow up was sufficiently long to enable outcomes to occur. |
| | Imprecision | Wide confidence interval in individual studies but narrow confidence interval for overall effect size (no effect) |
| | Inconsistency | Heterogeneity not explained by variability in the factors assessed |
| | Indirectness | No serious indirectness. All patients had IPF according to consensus criteria and were untreated. MMP-7 was measured at baseline in all studies, and 12-month mortality measured from IPD. |
| | Publication bias | No obvious funnel plot asymmetry, although unable to assess statistically due to small number of studies |
| | Certainty of evidence | Moderate certainty of evidence |

| | | |
|---|---|---|
| Disease progression (4 studies; 481 participants) | Risk of bias | All studies included well-defined participants diagnosed according to international consensus guidelines. Exposure and outcomes were measured objectively and consistently for all participants. Covariates were adjusted for using IPD. Follow up was sufficiently long to enable outcomes to occur. |
| | Imprecision | Wide confidence interval in individual studies but narrow confidence interval for overall effect size (no effect) |
| | Inconsistency | No significant heterogeneity |
| | Indirectness | No serious indirectness. All patients had IPF according to consensus criteria and were untreated. MMP-7 was measured at baseline in all studies, and overall mortality measured from IPD. |
| | Publication bias | No obvious funnel plot asymmetry, although unable to assess statistically due to small number of studies |
| | Certainty of evidence | High certainty of evidence |
| Change in FVC at 12 months (4 studies; 481 participants) | Risk of bias | All studies included well-defined participants diagnosed according to international consensus guidelines. Exposure was measured objectively and consistently for all participants. Change in FVC was measured objectively and consistently for all participants. Covariates were adjusted for using IPD. Follow up was sufficiently long to enable outcomes to occur. |
| | Imprecision | Wide confidence interval in individual studies but narrow confidence interval for overall effect size (no effect) |
| | Inconsistency | Inconsistency present across results from studies |
| | Indirectness | No serious indirectness. All patients had IPF according to consensus criteria and were untreated. MMP-7 was measured at baseline in all studies and change in FVC standardised using IPD. |
| | Publication bias | No obvious funnel plot asymmetry, although unable to assess statistically due to small number of studies |
| | Certainty of evidence | Moderate certainty of evidence. |

**Supplementary Table 8 –** GRADE (Grading of Recommendations, Assessment, Development and Evaluations) approach to rate the quality of evidence for the prognostic factor MMP-7
